# Practical evaluation of AI models to forecast a respiratory seasonal epidemic in Canada

**DOI:** 10.64898/2026.11.14.26368122

**Authors:** Benjamin Benteke Longaou, Warsame Yusuf, Monica-Gabriela Cojocaru, David Champredon

## Abstract

Public health organizations can benefit greatly from infectious disease epidemic forecasting to guide and support interventions and policies that reduce morbidity. However, accurate and reliable epidemic forecasting remains notoriously challenging and is an evolving field in which new scientific advances continue to support forecasting efforts. Computational models that underpin artificial intelligence (AI) models are promising developments that may improve seasonal epidemic forecasting. This study aims to compare the performance of a selection of AI models previously identified as well adapted to epidemic forecasting, including machine learning (XGBoost), deep learning models (Long Short-Term Memory (LSTM), Gated Recurrent Unit (GRU), Transformer), time-series foundation model (Chronos), and a hybrid model: physics-informed neural networks (PINNs), against statistical models (Autoregressive Integrated Moving Average (ARIMA), Prophet) and a compartmental epidemiological model (CEM) in seasonal epidemic forecasting across Canada.

The study uses weekly positivity-rate incidence of seasonal influenza A across eight Canadian provinces during the 2025/2026 season as a test case. Each model generated short-term (up to 5 weeks ahead) forecasts independently for each province from October 11, 2025, to March 28, 2026. We evaluated performance using mean absolute error (MAE) for each province.

Our results show that AI models, specifically GRU, Transformer, and Chronos models, achieve lower MAE than CEM in four of eight provinces (Alberta, British Columbia, Prince Edward Island, and Saskatchewan). Meanwhile, CEM and ARIMA outperform all AI models in three provinces (Ontario, Quebec, Manitoba) and one province (Nova Scotia), respectively. Furthermore, data pooling strategies that leverage multi-province data for the target virus or multi-virus data within individual provinces greatly improve deep learning models, and Chronos performs well under zero-shot forecasting in British Columbia. In contrast, Prophet always has the highest MAE, and PINN is worse than CEMs. Overall, AI models performed comparably to, or slightly better than, CEMs. However, these modest gains may not offset their lack of interpretability, high computational costs, and reliance on the specialized expertise required for implementation. We therefore conclude that AI models are best as a complement, rather than a replacement, to CEMs, which remain crucial for interpretability and informing decision-making.

## 1 Introduction

Seasonal acute respiratory infections impose a recurrent burden on healthcare systems and contribute substantially to annual morbidity [1]. At the onset of each season, surveillance systems track the emergence of epidemic activity, signaling the early phase of an outbreak. The progression of an epidemic is typically summarized using an epidemic curve, which represents the temporal evolution of indicators or targets such as incident cases, test positivity rates, hospital admissions, or infection-associated deaths. Whether strictly seasonal or driven by other transmission dynamics, such epidemics often follow a broadly predictable trajectory. In its simplest form, this trajectory consists of an initial phase of approximately exponential growth in incidence, followed by a peak as the pool of susceptible individuals becomes depleted, and subsequently a decline toward near-zero incidence. Despite this typical shape, it is usually challenging to predict the epidemic trajectory accurately. Therefore, the development of epidemic forecasting models is an active research area aiming to robustly forecast epidemic trajectories with satisfactory accuracy and confidence. Forecasting models are commonly designed to generate predictions ranging from a few weeks to several months ahead (depending on the disease and/or the goal of the forecasts), as well as to estimate peak magnitude and peak timing. As shown during multiple severe outbreaks, accurate forecasting models are essential for informing public health policy and operational planning [2]. Indeed, advance knowledge of epidemic trends enables policymakers to implement timely interventions, including situational awareness, vaccination campaign planning, expansion of healthcare capacity, and optimized resource allocation when peak timing and magnitude can be anticipated. In the absence of reliable forecasting models, decision-making and interventions can be suboptimal, leading to substantial human and financial costs.

Several methodological approaches have been proposed for epidemic forecasting, among which compartmental epidemiological models (CEMs) provide a mechanistic framework for describing disease transmission dynamics. CEMs are the most popular formulation, partitioning the population into relevant epidemiological states. The progression of the epidemic occurs through transitions of individuals between those epidemiological states, for example using systems of differential equations (the canonical Susceptible-Infectious-Recovered model [3]) or computational rules (agent based models). Machine learning algorithms underpinning “artificial intelligence” (AI) have had enormous interest from the scientific community over the past few years. Indeed, they have rapidly achieved capabilities that open new possibilities in terms of support and implementation of scientific projects across various fields. In particular, based on the promises of AI models, several groups have started to explore AI models to forecast epidemics [4, 5].

We may be at a pivotal moment where AI models are becoming essential tools for epidemic modellers, with the potential to transform how epidemic forecasting is conducted. In this study, we review traditional approaches to epidemic forecasting alongside recent advances in AI, with particular attention to their performance and applicability. Using a concrete case study of seasonal influenza forecasting in Canada, we assess the predictive accuracy of selected AI models and compare their performance with that of established epidemiological models. Importantly, our evaluation is grounded in a public health practice perspective. Rather than focusing on highly specialized or experimental AI developments, we assess models as they would be encountered by public health professionals, i.e., scientifically trained users who can engage with methods and implementation at a high level but are not AI specialists. Consequently, we focus on “ready-to-use” models, selecting those with accessible documentation, transparent methodologies, and user-friendly implementation. This approach reflects a pragmatic and implementation-oriented view, aiming to bridge the gap between methodological innovation and real-world deployment in public health settings.

Finally, we identify and discuss the practical challenges that AI models may introduce for public health organizations, including issues related to usability, interpretability, and integration into existing workflows.

The remainder of this paper is organized as follows. Section 2 provides background information and introduces the forecasting framework and baseline models considered in this study, including statistical, machine learning, deep learning, time series foundation, compartmental, and physics-informed neural network models. It also presents a literature scan of AI models for epidemic forecasting. Section 3 describes the study design, including the data and forecasting objective, training methodologies, data preparation and splitting, model training, and hyperparameter tuning. Section 4 presents the experimental results, focusing on point forecast performance, followed by a discussion of the practical implications for public health applications. Finally, Section 5 summarizes the main findings and discusses the practical challenges and considerations of using AI-based forecasting models in public health.

## 2 Background and baseline models

In this section, we briefly describe the forecasting process and models assessed in this study. Readers are invited to read the Supplemental Materials for more details.

### 2.1 Forecasting model

An epidemic curve can be represented as a time series, where observed incidence is periodically reported (e.g., daily, weekly, monthly, etc.). We denote the observed incidence sequence by *{y*_1_, *y*_2_, …, *y*_*T*_ *}*. The forecasting task consists of predicting the next *H* future observed incidences, namely *{y*_*T* +1_, …, *y*_*T* +*H*_*}*, where *H* and *T* represent the forecast horizon and total number of observations, respectively. Formally, the forecasting task can be expressed as fitting, or “learning”, a function *f* such that

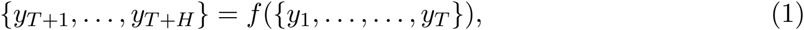

where *f* is called a forecasting model.

Forecasts for a given horizon may be either deterministic or probabilistic. Deterministic forecasts produce a single predicted value and are commonly referred to as *point forecasts*. In contrast, *probabilistic forecasts* provide a predictive distribution or a set of quantiles, thereby characterizing uncertainty in future outcomes (Figure 1).

**Figure 1.**
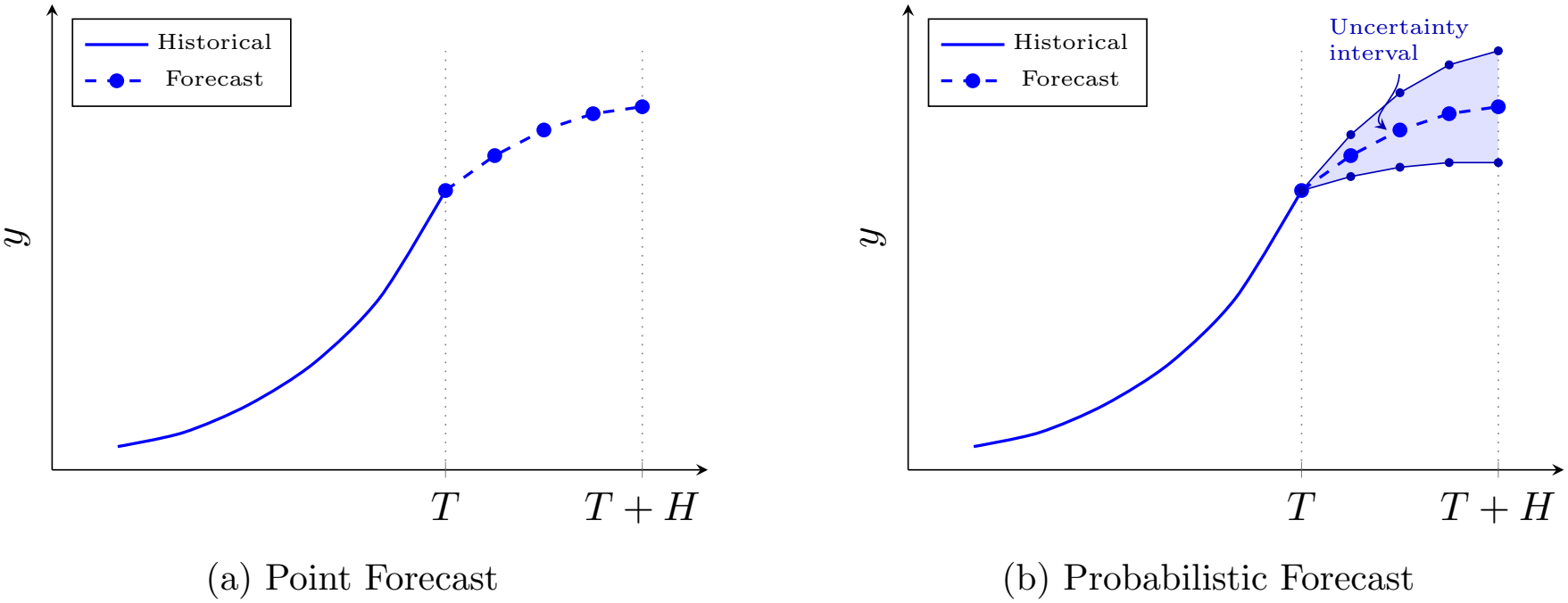
Comparison of point vs. probabilistic forecasts.

Ideally, forecasts should be accurate, stable, and, in the probabilistic setting, accompanied by well-calibrated and sufficiently narrow uncertainty intervals.

In this paper, we consider a diverse set of forecasting models. Specifically, we consider compartmental mechanistic models (CEMs), statistical models, machine learning (ML) models, deep learning (DL) models, physics-informed neural networks (PINNs), and time-series foundation models (TSFMs). Furthermore, in order to compare all evaluated models, our primary focus is centered strictly on point forecasts as some models cannot natively generate probabilistic forecasts.

Before describing the individual models, we first explain how we transform a time series into training samples for models that require an explicit supervised-learning formulation. Standard ML and DL models are inherently static; they expect tabular data with distinct feature vectors (*X*) and target vectors (*y*) and cannot process raw, continuous sequence data. To bridge this gap, we use *lag embedding* (the sliding-window approach). This approach converts a single long time series into training supervised samples (input-target pairs), where the feature vector consists of a window of *p* past observations and the prediction target is the next *h* future observations. By framing temporal dependencies as static tabular features, lag embedding lets us cast sequence forecasting as a standard regression task and leverage powerful ML and DL algorithms.

### 2.2 Lag embedding

Lag embedding, also called training sample creation, transforms a univariate time series into a supervised learning problem by using a sliding window of past observations as predictors and subsequent observations as targets. For a given time series, a specified number of past observations form each input sample, while the following observation(s) form the corresponding forecasting target. Moving the window through the time series generates a sequence of input-target pairs for model training.

This representation enables ML and DL models to learn temporal dependencies and patterns from historical observations. The resulting samples are subsequently used to train the forecasting models.

### Recursive forecasting strategy

Recursive forecasting produces multi-step-ahead forecasts sequentially, one step at a time. The model predicts the next observation based on the latest available observations. The model then re-injects the predicted value into the input sequence and uses it, along with the remaining recent observations, to generate the next prediction. This is repeated recursively until the desired forecasting horizon is reached. Thus, predictions from previous steps are used as input for subsequent forecasting steps.

Consider a univariate time series with 10 observations: *y* = [*y*_1_, *y*_2_, *y*_3_, *y*_4_, *y*_5_, *y*_6_, *y*_7_, *y*_8_, *y*_9_, *y*_10_].

We construct supervised learning data using a lag order (window size or context length) *p* = 3, where each input *x*_*i*_ consists of the previous three values of the series, and the target *y*_*i*_ is the next value. Thus, the time series is transformed into a supervised training dataset:

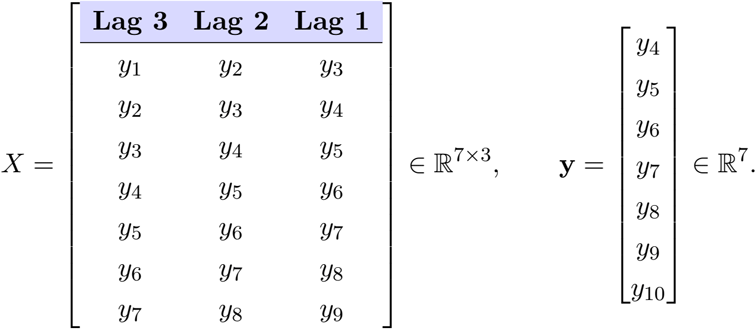

For models requiring lag embedding, the number of training samples generated from the original time series is *T − p*. In the example above, with *T* = 10 and *p* = 3, this yields 10 *−* 3 = 7 training samples. This formulation provides a unified framework for predicting future incidence values from historical epidemic data.

Once the model is trained, it can be used for forecasting. Assume that the last available input window consists of the observations [*y*_8_, *y*_9_, *y*_10_]. For a forecasting horizon of *H* = 5, recursive forecasting proceeds by iteratively feeding predictions back into the model:

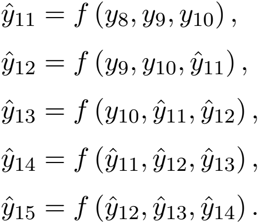

In this approach, each predicted value becomes input for the next prediction step. In the example above, we illustrate *recursive forecasting*, in which a single one-step-ahead model is iteratively applied to generate multi-step forecasts. Some forecasting strategies instead train a separate model for each prediction horizon; this approach is known as *direct forecasting*. In contrast, *multi-output forecasting* uses a single model to generate predictions for multiple future horizons simultaneously. Some strategies also combine two or more of these approaches and are referred to as *hybrid forecasting*.

While ML and DL models require explicit lag embedding to construct input training samples, the statistical models and TSFMs handle lag-window construction and recursive multi-step forecasting internally. Consequently, these architectures omit these preprocessing steps during feature engineering. In contrast, CEMs and PINNs do not rely on lag embeddings or recursive forecasting.

In this study, we consider *p ∈ {*2, 4, 8, 16*}* for models requiring sample creation through lag embedding. AI and statistical models use a recursive strategy and generate one-step-ahead forecasts.

### 2.3 Learning a forecasting model

Given a dataset of training samples 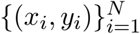 extracted from a time series via lag-embedding, let *f*_*θ*_ denote the forecasting model parameterized by a vector of parameters *θ*. The goal of learning is to find the optimal parameter set 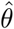 that minimizes a given loss function *ℒ* over the training instances:

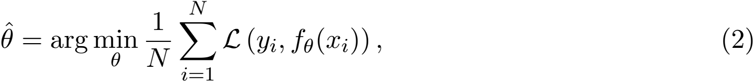

where *y*_*i*_ represents the ground-truth value and *f*_*θ*_(*x*_*i*_) is the model’s prediction.

For a *point forecasting* task, the prediction is a single scalar *ŷ*_*i*_ = *f*_*θ*_(*x*_*i*_), and the objective typically minimizes a point error loss function, such as the Mean Squared Error (MSE):

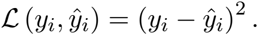

This framework is primarily intended for ML and DL models, while Section 3 describes the training procedure for each model separately.

### 2.4 Compartmental mechanistic models

Compartmental epidemiological models (CEMs) are the standard mechanistic models that are widely used for infectious disease modelling, and have their origin in the 1920s with the work of Kermack and McKendrick [6]. The fundamental features of these models are they i) compartment individuals of the population into epidemiological states, and ii) describe mathematically the migration dynamics of individuals between the compartments as the infections spread in the population. The temporal evolution of compartment sizes is typically modelled using systems of ordinary differential equations (ODEs) that describe transition rates between compartments, or equivalently, renewal equations [7]. One of the simplest compartmental models to simulate epidemics is the Susceptible-Infectious-Recovered (“SIR”) model. Individuals who have never been infected (i.e., still susceptible to infections) are grouped in the “susceptible” compartment, and the size of this compartment is noted *S*. Infectious individuals, their number is noted *I*, are grouped in the “infectious” compartment. Finally, the “recovered” compartment (R) is filled with individuals who are no longer infectious and immune for life. The canonical modeling of the transition rates of this simple SIR model is given by the following system of ODEs

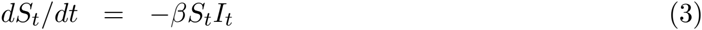

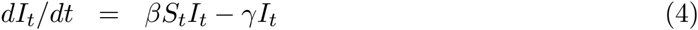

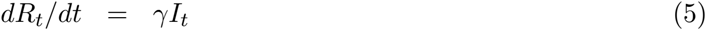

where *β* and *γ* represent the contact and recovery rates, respectively.

The CEMs can be designed to represent very complex epidemiological processes. The practical limit to their complexity and size being the available data to fit the model parameters to and computational power (most CEMs are not analytically tractable and require numerical solvers).

CEMs are scientifically grounded as they mathematically represent biological and epidemiological processes underlying disease transmission and natural history, behaviors, and interventions. These models are inherently interpretable and allow the estimation of key epidemiological parameters (e.g., basic reproduction number *R*_0_). They can also be used to assess the potential impact of public health interventions or behavioral changes on epidemic dynamics.

CEMs are often used to forecast infectious disease epidemics because they provide an attractive balance between modeling tractability and representativeness of the real world. Forecasting is conceptually straightforward: once the model is fitted to observations, epidemic trajectories are simulated forward in time (beyond the last observation date) up to a predefined time horizon, constituting the forecasted epidemic.

The CEM used in this study is a simple SIR model implemented with a renewal equation, presented in Subsection 3.3.

### 2.5 Statistical models

Many statistical models [8, 9] have been developed for time-series forecasting, and reviewing them is beyond the scope of this study. Statistical models are data-driven models that learn patterns and relationships from historical data to predict future outcomes. These models are often used for forecasting because they can capture complex temporal dependencies and trends without requiring explicit mechanistic assumptions about the underlying processes. Researchers have applied statistical models to forecast various targets in infectious disease epidemics [10, 11, 12, 13, 14].

Many statistical models have been developed for time series forecasting, including autore-gressive integrated moving average models (ARIMA), seasonal autoregressive integrated moving average (SARIMA), exponential smoothing (Holt–Winters), and Prophet. In this study, we selected ARIMA [15] and Prophet [16] as representative statistical forecasting models. ARIMA has been shown to be effective for short-term forecasting when temporal patterns are approximately linear and stationary after differencing. Its interpretability and relatively low computational complexity have contributed to its extensive use in epidemiological forecasting and disease surveillance.

Many studies have proved the effectiveness of ARIMA in infectious disease forecasting. ARIMA has been used to forecast the spread of COVID-19 in Italy [17], estimate the number of confirmed and recovered COVID-19 cases [18], and predict trends in COVID-19 prevalence in Italy, France, and Spain [19]. ARIMA has also been applied to forecast influenza in China beyond COVID-19 [20]. The authors found that ARIMA performed well, especially for short-term epidemic forecasting in modeling temporal dynamics in public health data.

The second statistical forecasting model selected for our study Prophet [16], as previous studies report competitive forecasting accuracy, especially for time series with long training histories [21]. It has also been used in epidemiological forecasting, e.g., for influenza prediction [20]. Its inclusion therefore provides a complementary statistical approach to ARIMA for evaluating forecast performance across different temporal patterns and forecast horizons.

More broadly, the distinction between traditional statistical forecasting models and CEMs is not always definitive in terms of predictive performance. Previous studies have shown that these approaches may outperform one another depending on the forecasting scenario and disease dynamics [22]. For example, collaborative forecasting studies on influenza-like illness demonstrated that statistical forecasting models can achieve competitive accuracy in epidemic prediction tasks [23].

### 2.6 AI models

Given that the primary audience for this report is public health practitioners, many of whom may have limited familiarity with AI models, this section offers a concise, high-level description of how they work.

*AI models* is a loose, umbrella term that generally refers to any computational model used to perform tasks associated with artificial intelligence, such as prediction, classification, generation, or decision-making. Here, leaning on recent literature, we focus on modern models relevant to forecasting infectious disease epidemics.

In this paper, we focus on two categories of AI models: machine learning (ML) and deep learning (DL). *Machine learning* is a class of statistical and computational methods that learn patterns or relationships from data to make predictions, classifications, or decisions, without being explicitly programmed with fixed rules [24]. Learning types are typically categorized by the amount of human/expert guidance provided. In traditional machine learning, the three main types of learning are supervised, unsupervised, and reinforcement learning. *Supervised learning* is a form of machine learning in which a model is trained on labeled data, that is, pairs of inputs and known outputs, to learn a mapping that can predict the output for new, unseen inputs. In *unsupervised learning*, the model receives only unlabelled inputs and discovers patterns or structures, such as clustering similar data points, detecting anomalies, or reducing the dimensionality of complex datasets while preserving key information. In this paper, we focus on supervised learning, specifically the regression task, as our goal is to forecast continuous variables (e.g., influenza incidence) rather than predict categorical labels.

### 2.1 Machine learning models

Machine learning models have been widely applied to epidemic forecasting [25], with various models used for different forecasting tasks. Linear regression and its regularized versions (Ridge, LASSO, and Elastic Net), decision trees, support vector machines (SVMs), and tree-based ensemble methods [26, 27] such as random forest and extreme gradient boosting (XGBoost) are commonly used in epidemic forecasting. In this paper, we use XGBoost as a representative decision-tree model, given its widespread use and strong performance in various forecasting tasks [20, 28, 29].

*Decision tree regression* is a non-parametric supervised learning method that models nonlinear relationships between input features and a continuous target variable. The model recursively partitions the feature space into disjoint regions by selecting split points that minimize prediction error, typically measured using Mean Squared Error (MSE). At each terminal node (leaf), the predicted output is the mean value of the training samples in that region. Figure 2 illustrates how a decision tree works.

**Figure 2.**
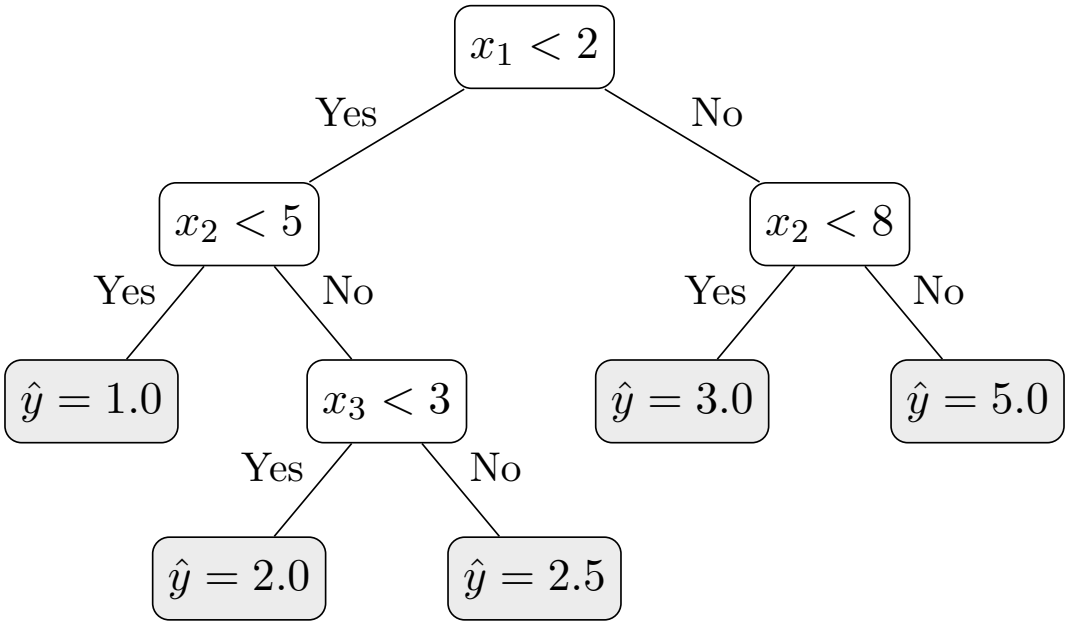
Illustration of a decision tree for predicting the target variable from the input features (*x*_1_, *x*_2_, *x*_3_)

Decision tree models are easy to understand and interpret, but have high variance and tend to overfit the training data, especially in complex datasets [30]. To overcome this limitation, tree-based ensemble methods such as Random Forest and XGBoost combine multiple decision tree models to improve the predictive performance and robustness.

#### XGBoost

XGBoost is a widely used tree-based ensemble method for classification and regression on tabular data, known for its strong predictive performance and ability to model complex relationships in structured datasets [29]. It efficiently implements gradient-boosted decision trees (GBDT) by combining multiple weak decision trees into a strong predictive model. Given a set of training observations 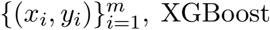 builds an ensemble of *n* decision trees and gives the prediction

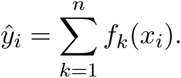

The *k*-th decision tree is indicated by *f*_*k*_. Figure 3 shows the basic structure of XGBoost. Unlike bagging-based ensemble methods like Random Forest, XGBoost constructs trees sequentially in an additive way. Each subsequent tree is trained to correct the current ensemble’s errors, enabling the model to gradually reduce prediction error. The higher predictive accuracy and improved generalization capabilities of XGBoost are obtained by combining the predictions of several trees via this boosting procedure.

**Figure 3.**
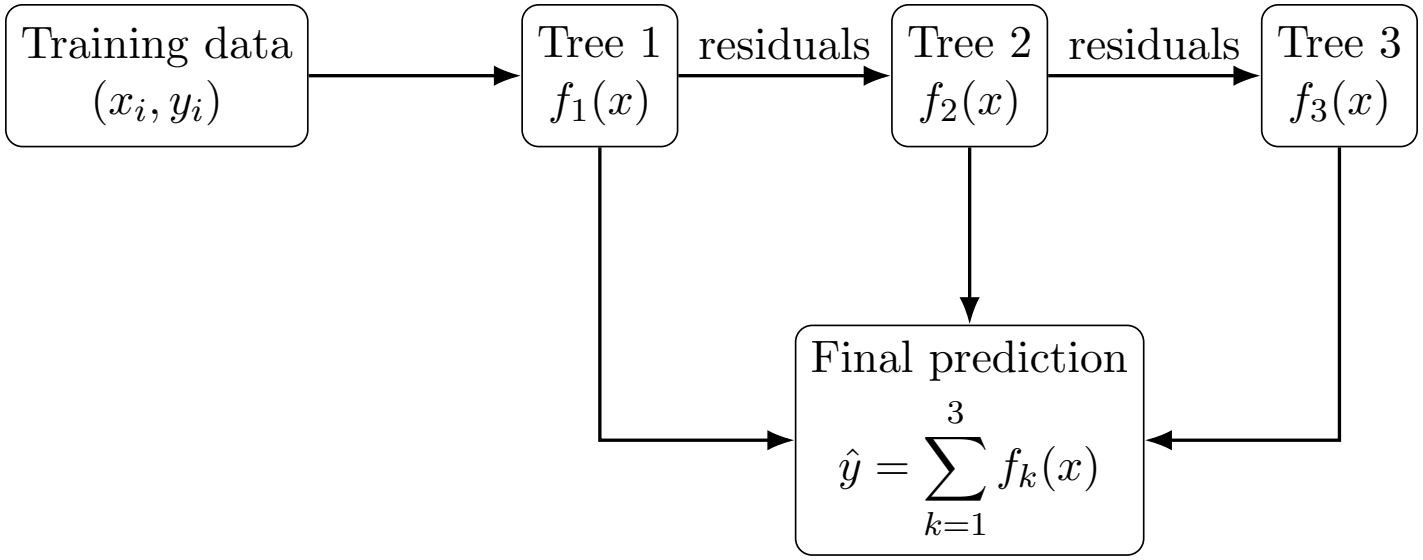
Illustration of XGBoost, showing the sequential construction and combination of decision trees to produce the final prediction.

##### Example 2.1.

*Here is a very simplistic example illustrating the basic concepts underpinning XGBoost in the context of epidemic forecasting. The goal is to predict next week’s cases using only previous weeks’ cases. Suppose the historical data are:*

*Note that, because XGBoost is a “supervised” method, the data in Table 1 is “labeled” where the input are the cases from the past three weeks and outputs are cases for the next week. In this setting, XGBoost learns a collection of small decision trees that capture simple patterns in the epidemic trajectory using only past case counts. For example, Tree 1 may encode a rule such as: “if the number of cases in the previous week exceeds 100, then predict an increase in cases”. Tree 2 may capture acceleration, for instance: “if the number of cases last week is much higher than two weeks ago, then add an additional increase to the prediction”. Tree 3 may represent sustained growth, such as: if cases have been increasing over the last three weeks, then further amplify the forecast. Each tree models a simple aspect of epidemic dynamics*.

*To make a forecast, the current observations, say 110 cases last week, 80 two weeks ago, and 60 three weeks ago, are passed through all the trees. Each tree contributes a small numerical value (for example, Tree 1 predicts, say*, +100, *Tree 2 predicts* +20, *Tree 3 predicts* +10*), and the final prediction for the cases next week is obtained by summing contributions from all the trees (e*.*g*., *130 cases for the next week in this simplistic example). In this way, the model combines multiple simple rules derived from past epidemic behavior to produce an overall forecast*.

Note that for real-world applications, more than hundreds of trees are generated, making the interpretation of the “discovered” rules by humans practically impossible (that is, outputs results are treated as a “black box”).

To summarize simply, XGBoost forecasts an epidemic by learning how past case trajectories evolve, using many small decision trees that capture key patterns (like growth and acceleration). XGBoost has been chosen on the basis of evidence from earlier forecasting studies. As shown in [31], XGBoost outperformed deep learning models, such as Recurrent Neural Networks (RNNs) and Long Short-Term Memory (LSTM) networks, for tabular COVID-19 forecasting at 1 to 5 weeks ahead. Similarly, [32] found that Gradient Boosting Regression Trees (GBRTs) can outperform state-of-the-art deep neural network (DNN) models by efficiently engineering input and output structures. This means XGBoost should serve as the tree-based machine learning baseline in our study.

### 2.2 Deep learning models

Deep learning (DL) is a type of machine learning that uses artificial neural networks with multiple layers to learn hierarchical data representations, allowing complex relationships and patterns to be modeled. The Deep Neural Network (DNN) models [8, 12, 33, 34, 35, 36] employed in epidemic (short-term) forecasting are simple Recurrent Neural Networks (RNNs), Long Short-Term Memory (LSTM), Gated Recurrent Unit (GRU), BiLSTM, BiGRU, and transformers.

#### Artificial Neural Networks

An Artificial Neural Network (ANN) is a deep learning technique based on biological neural systems and comprises neurons connected and arranged in layers; it learns the relationship between inputs and outputs by modifying its weights and biases during training. The neurons are the fundamental units of computation in an ANN. Each one calculates a weighted sum of its inputs, adds a bias term, and then applies a nonlinear activation function. Activation functions let the network model complex relationships by introducing non-linearity; without them, the network would function as a linear one no matter how deep it is. An ANN consists of input, hidden (optional), and output layers: the input layer takes in the data, the hidden layers learn intermediate representations, and the output layer generates predictions.

#### Feedforward

A feedforward neural network is a network in which information flows in one direction through fully connected layers.

#### Multi-layer perceptron

A multi-layer perceptron (MLP) is a common example of a feedforward neural network with one or more hidden layers and non-linear activations.

#### Deep neural network

A deep neural network (DNN) is an MLP with many hidden layers to learn progressively more abstract patterns. DNNs can learn complex nonlinear relationships and hierarchical representations from high-dimensional data. Depending on the architecture, they can also capture temporal dependencies over different time scales.

In time series forecasting, these models act as static regressors, i.e., using a fixed window of past observations to predict future values, which means they lack memory and struggle to capture long-term temporal dependencies. As a result, they are less suited to sequential data; hence, *recurrent neural networks* (RNNs) and their variants were introduced to incorporate memory by allowing time-dependent hidden states to accumulate information over time.

##### Example 2.2.

*In this setting, a deep neural network learns a hierarchy of hidden representations from past epidemic data through multiple layers of neurons. Assume the first layer learned that it needed to compute a simple weighted combinations to represent an overall level by applying weights of, say, 0*.*5, 0*.*3 and 0*.*2 to the number of cases of 1, 2 and 3 weeks ago respectively. This first layer also needs to represent the recent increase signal by calculating the difference between the number of cases 1 and 2 weeks ago. Assume the second layer can then combine these intermediate quantities, for instance detecting growth by amplifying the increase signal by a factor of, say, 1*.*2, and summing it with the overall level to produce an aggregate growth indicator. A third layer may further transform this into a higher-level representation such as outbreak momentum, for example by scaling this signal by, say, 1*.*1. These transformations are not explicit rules (unlike XGBoost) but learned numerical mappings*.

*Let’s take again the data in Table 1 as a simplistic example. Given inputs of 110 cases last week, 80 two weeks ago, and 60 three weeks ago, to make a forecast the input values are propagated through all three layers: the first layer extracts basic signals (*0.5 *×* 110 + 0.3 *×* 80 + 0.2 *×* 60 = 91*) and increase of 110 - 80 = 30. The second layer calculates the intermediary value* 91 + 1.2 *×* 30 = 127. *Finally, the third layer interprets this as high epidemic momentum and multiply the number calculated in the second layer by 1*.*1, yielding* 1.1 *×* 127 *≈* 140. *As the final output layer this representation provides the numerical prediction of approximately 140 cases for the next week*.

Put simply, an artificial neural network uses successive layers to progressively transform raw inputs into more abstract quantities that directly inform the forecast.

Note that in practice, a large number of layers is often used, making the interpretation of hundreds, if not millions, of weights impossible by humans, who must consider this whole forecasting process as a “black box” (like for XGBoost). See Appendix A for a more detailed description of the various types of neural networks.

#### Long Short-Term Memory

Long Short-Term Memory (LSTM) networks [37] are a type of RNN designed to overcome the “vanishing gradient problem” (i.e., when the network forgets how to learn from earlier information, because the updates become too tiny to have any effect) and capture long-range dependencies in sequential data. They maintain two forms of memory, a long-term and a short-term, and use gating mechanisms to control what information is kept, updated, or discarded over time.

#### Gated Recurrent Units

Gated Recurrent Units (GRUs) [38] are a simpler version of LSTMs designed to handle the vanishing gradient problem in RNNs. They use fewer gates (mainly an update gate and a reset gate) to control memory, making them faster and simpler while still capturing long-term patterns in sequential data. Appendix A provides additional architectural details.

#### Transformers

The transformer architecture, introduced in [39], is a neural network architecture designed to handle sequential data efficiently that relies on “self-attention” mechanisms. Transformers have revolutionized natural language processing and have been adapted for time series forecasting because they can model long-range dependencies and parallelize computations effectively. The core of a Transformer is self-attention, which lets the model decide how important each time step is when making a prediction. It does this by considering all the data at once, capturing the most important parts of the information, assigning weights to all inputs, and combining them into a contextual representation. An encoder processes the input sequence, and a decoder uses this information to produce the forecast, while multi-head attention captures different types of relationships in the data simultaneously, making it particularly effective for complex time series forecasting tasks where multiple factors may influence the target variable.

### 2.7 Time Series Foundation Models

When neural networks are used for forecasting infectious diseases, they usually require large amounts of data, can be prone to spurious correlations, and may not perform well when applied beyond the conditions found in the training data [40]. In this context, so-called Time Series Foundation Models (TSFMs) [41, 42] have emerged as a promising solution, shifting from task-specific forecasting models to unified models that can generalize across fields such as epidemiology, finance, energy, and climate. Like Large Language Models (LLMs) in natural language processing, TSFMs are pre-trained on large, diverse sets of time series data, which may reduce the need for extensive task-specific training.

TSFMs are trained on datasets that differ greatly in sequence length and temporal resolution, covering observations on a quarterly, monthly, weekly, daily, and hourly basis. The datasets can contain real-world, synthetic, or hybrid time series drawn from a variety of domains as mentioned above. Through pretraining, TSFMs learn general temporal patterns, such as trends, seasonality, and long-range dependencies, which can then be applied to downstream tasks or previously unseen datasets.

As illustrated in Figure 4, TSFMs generally operate through two stages: *pretraining* and *adaptation*. During pretraining, researchers define a model architecture and train it on diverse time series using supervised or self-supervised learning. Most TSFMs use transformer-based architectures, including encoder-only, decoder-only, and encoder-decoder models, although non-Transformer architectures. During adaptation, the pretrained model is tailored to a specific forecasting task or dataset to leverage the temporal representations learned during pretraining. Common adaptation strategies include zero-shot inference, fine-tuning, prompt engineering, and tokenization-based [42].

**Figure 4.**
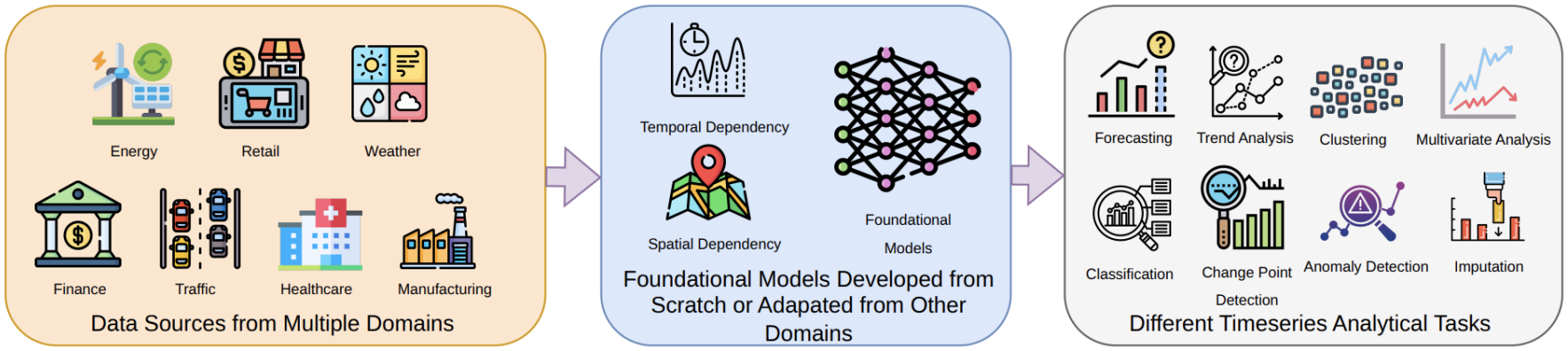
Pretraining Time Series Foundation Models on diverse data sources for downstream generalization [41].

**Figure 5.**
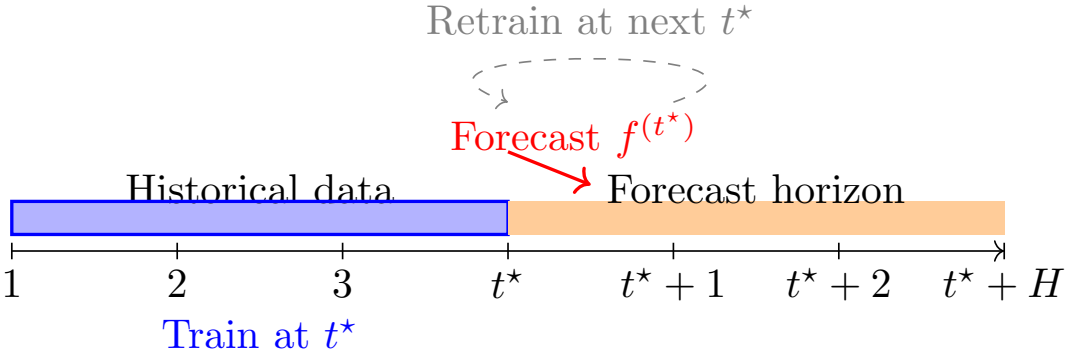
Rolling-origin forecasting procedure with model retraining at each as-of date *t*^*⋆*^ and generation of *H* consecutive out-of-sample forecasts.

We examine two adaptation methods: zero-shot and fine-tuning. With zero-shot inference, we use the pre-trained model directly on the target dataset without further training, enabling us to assess its ability to generalize to previously unseen datasets. This method is especially appropriate when historical data are scarce and offers a ready-made forecasting approach for various diseases, regions, and outcomes. In fine-tuning, the pre-trained model is modified to suit the specific forecasting task by updating its parameters with task-specific data. This fine-tuning process can involve updating all of the model’s parameters or only some, leaving the rest unchanged. In this study, we consider both parameter-efficient fine-tuning and full fine-tuning.

Recently released lightweight, open-source versions have made it increasingly feasible to put TSFMs into practical use, as their effectiveness has been shown in specific applications such as epidemic forecasting [21, 43, 44, 45].

#### Chronos model

For this study, we choose the Chronos model [46] as an example of a TSFM because of its comprehensive documentation and accessible off-the-shelf Python implementation. This makes Chronos usable for users without specialist expertise in TSFMs. Moreover, Chronos and TimesFM [47] have shown higher accuracy than other TSFMs [44]. Chronos treats numerical time-series forecasting as a sequence prediction task, similar to language modeling. Each time series is normalized and then binned into a sequence of discrete tokens. This allows diverse time series to be represented in a unified token space. These token sequences are input to a pretrained model trained to predict future tokens based on past ones. Finally, we assess Chronos using zero-shot and fine-tuning.

### 2.8 Physics-informed neural networks

Physics-informed neural networks (PINNs) are neural networks that are trained not only on data, but also guided by known physical laws (like equations describing how a system behaves). In simple terms, they learn from data while being constrained within the known rules of physics, so their predictions stay realistic (at least in the realm of what is physically possible). In the context of epidemic models, using standard SIR model, the constraints can be driven by the dynamics of the subgroup population sizes (e.g., susceptible, infectious, recovered, etc.) and initial conditions. For a more detailed discussion of PINNs and their applications to epidemic modeling and forecasting, we refer the reader to [48, 49, 50, 51, 52, 53, 54].

### 2.9 Literature scan on AI models to forecast epidemics

To provide context, we conducted a broad, non-systematic literature review to highlight published studies that used AI models to forecast epidemics. Several recent reviews and broader perspectives have examined the use of artificial intelligence and machine learning in infectious disease modeling, epidemic forecasting, disease surveillance, and pandemic preparedness [4, 55, 56, 57, 58, 59, 60, 61, 62]. Recent work has also specifically reviewed the application of AI to influenza forecasting [40].

We focus on *respiratory* diseases in this review because our study uses influenza A as a working example. We do not review the studies using “traditional” statistical and compartmental mechanistic models, as they are too numerous and such a review would be beyond the scope of this study.

ML models can identify relationships between input variables and desired outcomes and have become increasingly used in epidemic forecasting. For instance, in [20], XGBoost, seasonal autoregressive integrated moving average (SARIMA), Prophet, and Holt–Winters were compared for forecasting monthly influenza cases in Fuzhou. XGBoost outperformed the statistical models in both fitting and forecasting, and Prophet also showed competitive predictive performance. Although the Holt–Winters additive model fitted the data less well than SARIMA, its forecasts were more accurate. Similarly, [63] found that a support vector regressor (SVR) performed better than ARIMA in forecasting cumulative COVID-19 cases six days ahead for 10 Brazilian states. These studies show that ML models can provide competitive epidemic forecasts by relying on data-driven relationships between predictors and outcomes.

DL models have likewise shown powerful predictive abilities by learning the complex relationships between input variables and the target outcomes. After having been trained, they are able to use observed data to predict future values, which has made them ever more useful for forecasting epidemics. A number of DNN architectures have been used in short-term epidemic forecasting, such as RNNs, LSTM, GRUs, bidirectional LSTMs (BiLSTMs), bidirectional GRUs (BiGRUs), and Transformer models [8, 12, 33, 34, 35, 36].

LSTM models have been extensively studied for epidemic forecasting because they can model temporal dependencies. They have been used for forecasting the transmission of COVID-19 in Canada [64] as well as for short-term influenza forecasting [65]. Recently, [66] showed that LSTM-based models are effective for short-term influenza forecasting and mentioned the importance of including meteorological variables and national public holidays in the predictive framework.

Researchers have also investigated the use of transformer-based models for forecasting epidemics, since their attention mechanism can model complex temporal relationships and dependencies over time. For instance, Transformer models have been applied to predict influenza prevalence [67]. In a comparative study involving short-term forecasting of Influenza A, [68] found that the Transformer model performed better than LSTM, GRU, BiLSTM, and ARIMA. Likewise, [69] compared various models for short-term Influenza A forecasting and found that deep learning models outperformed traditional statistical models such as ARIMA and exponential smoothing (ETS), with the Transformer model achieving the best results. Further comparative studies have also examined LSTM, GRU, and BiLSTM models against ARIMA, and in those studies LSTM showed competitive predictive performance [70].

More recently, TSFMs are presented as a promising model family, leveraging massive pre-training on diverse sequential data to achieve zero-shot forecasting. Recent assessments of TSFM architectures have revealed various operational trade-offs [21]. Models like TabPFN [71], Chronos, TimesFM, and TimeGPT [72] tend to perform well not only for short- and long-term point forecasting, but also for peak-period forecasting, where TimesFM performs well. Notably, [44] showed that Chronos and TimesFM deliver superior overall accuracy compared to other TSFMs, directly motivating our interest in evaluating Chronos’s performance dynamics for public health forecasting.

The strong performance of deep learning models also raises the question of whether complex DL architectures are always necessary for time series forecasting. Ensemble learning methods, particularly tree-based boosting models, provide an alternative data-driven approach. For example, Gradient Boosting Regression Trees (GBRTs) can compete with, and in some cases outperform, state-of-the-art DNN models through careful engineering of the input and output structures [32]. These findings suggest that effective feature engineering combined with ensemble learning can provide competitive forecasting performance without the complexity of deep neural networks.

Recently, researchers have explored physics-informed neural networks (PINNs) as an extension of DNN models. PINNs use epidemiological knowledge or governing equations into the learning process, for instance, an SIR model, so their predictions are guided by known disease dynamics rather than relying solely on data. According to [52], PINNs outperformed conventional DNN models for epidemic forecasting, highlighting the advantage of combining data-driven learning with domain knowledge.

[61] reviews and compares traditional statistical and deep learning methods for epidemic forecasting.

This literature scan suggests that AI models, including ML models, DL models, and PINNs, can have competitive forecasting accuracy. For these reasons, we include all these models. First, performance varies. First, performance varies by disease, forecasting horizon, and region. Second, complex models do not always increase forecasting accuracy; for instance, XGBoost can perform similarly to more complex deep neural network architectures. Finally, PINNs are hybrid methods that combine DNNs and CEMs; they sometimes outperform DNNs.

### 2.10 Limitations of AI models

Although AI and machine learning models have been successfully applied to infectious disease forecasting, significant limitations continue to constrain their practical implementation.

First, interpretability remains a major challenge. These models can produce accurate forecasts of disease dynamics but often provide limited insight into the mechanisms underlying their predictions, as they lack explicit mechanisms to depict how disease progression actually occurs. This lack of transparency matters in epidemiological applications, where forecasts must be interpretable and actionable for policymakers and epidemiologists. Moreover, purely data-driven models are not inherently grounded in epidemiological principles and therefore cannot directly provide scientifically meaningful quantities, such as the transmission rate or the basic reproduction number.

Second, their predictive performance and robustness depend heavily on data quality and availability. This drawback is relevant for disease data, which are often sparse, under-reported, delayed, noisy, and over-dispersed. Consequently, their performance may deteriorate, as is often the case for emerging or localized outbreaks. AI models can learn spurious correlations rather than meaningful relationships, which may limit their ability to generalize to new outbreaks.

Finally, practical implementation presents significant operational hurdles. Training and deploying these traditional AI models can be computationally expensive, requiring extensive hyperparameter tuning across high-dimensional parameter spaces. Furthermore, successful implementation demands specialized technical expertise in machine learning engineering, and these architectures can struggle with long-term forecasting.

## 3 Study design

This study aims to assess the both the practicality and performance of selected AI models to forecast influenza A in the most populous provinces in Canada. The choice for influenza A is motivated by the fact that this is a seasonal infection which has been part of the PHAC’s respiratory viruses surveillance system for more than about two decades, hence there is a lot of data that can be leveraged by the models. Although influenza A has, nearly every season, the expected typical shape of an epidemic – exponential growth, a single peak, followed by a exponential decrease – the start of seasonal epidemics as well as their peak are variable from season to season, making it a challenging epidemic to forecast. Moreover, seasonal influenza A is epidemiologically important because of its burden on health systems and global morbidity. To summarize, seasonal epidemics of influenza A in Canada provide a good balance (for our study) between data quality, data quantity, season-to-season variability and public health relevance.

### 3.1 Data and goal

We used the positivity rate collected by PHAC’s national respiratory surveillance system. Number of laboratory tests performed and number of tests confirmed positive for acute respiratory infections are reported weekly at the provincial level. Data is publicly available at https://www.canada.ca/en/public-health/services/surveillance/respiratory-virus-detections-canada.html. The positivity rate controls for the testing effort, which may not be constant throughout the season. We extracted historical weekly surveillance data for laboratory-confirmed influenza A cases across eight Canadian provinces: British Columbia (BC), Alberta (AB), Saskatchewan (SK), Manitoba (MB), Ontario (ON), Québec (QC), Nova Scotia (NS), and Prince Edward Island (PE). Model training utilizes historical time series starting from the 2006–2007 influenza season. The primary evaluation period spans the 2025–2026 season

The goal was to forecast the 2025/26 influenza A season independently for each selected province. Forecasts were generated on a rolling weekly basis, with the first as-of date October 4, 2025. At each subsequent as-of date, the newly available observation was added to the training/fitting dataset, and the models were refitted before generating the 1 to 5 weeks next forecasts.

This rolling procedure continued through March 28, 2026, which was the last data point considered

In the following subsections, for each forecasting model considered, we describe its model configuration, hyperparameter search space and tuning procedure, training procedure, and forecast generation strategy.

### 3.2 Training methodologies

Starting from an as-of date *t*^*⋆*^, where we use all information available up to and including *t*^*⋆*^, we train the forecasting model and generate forecasts for the next *H* steps. Thus, for each as-of date *t*^*⋆*^, the model is trained using the available data and produces *H* consecutive out-of-sample forecasts, 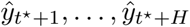 . The as-of date then advances through the fixed rolling test window, and the procedure repeats. All model fitting and preprocessing at each as-of date use only information available up to that date, thereby preventing look-ahead bias and ensuring strict temporal consistency.

To evaluate the impact of information sharing across geographical regions and viral types (COVID-19 and respiratory syncytial virus (RSV)), we train the selected forecasting models using four distinct paradigms (*local, global A, global B*, and *partial global*, summarized in Table 2).

**Table 1:** Hypothetical data used in an illustrative example for machine learning models.

| **Week** | **Cases last week** | **Cases 2 weeks ago** | **Cases 3 weeks ago** | **Prediction target** |
|---|---|---|---|---|
| 4 | 70 | 55 | 50 | 90 |
| 5 | 90 | 70 | 55 | 120 |
| 6 | 120 | 90 | 70 | 150 |

**Table 2:**
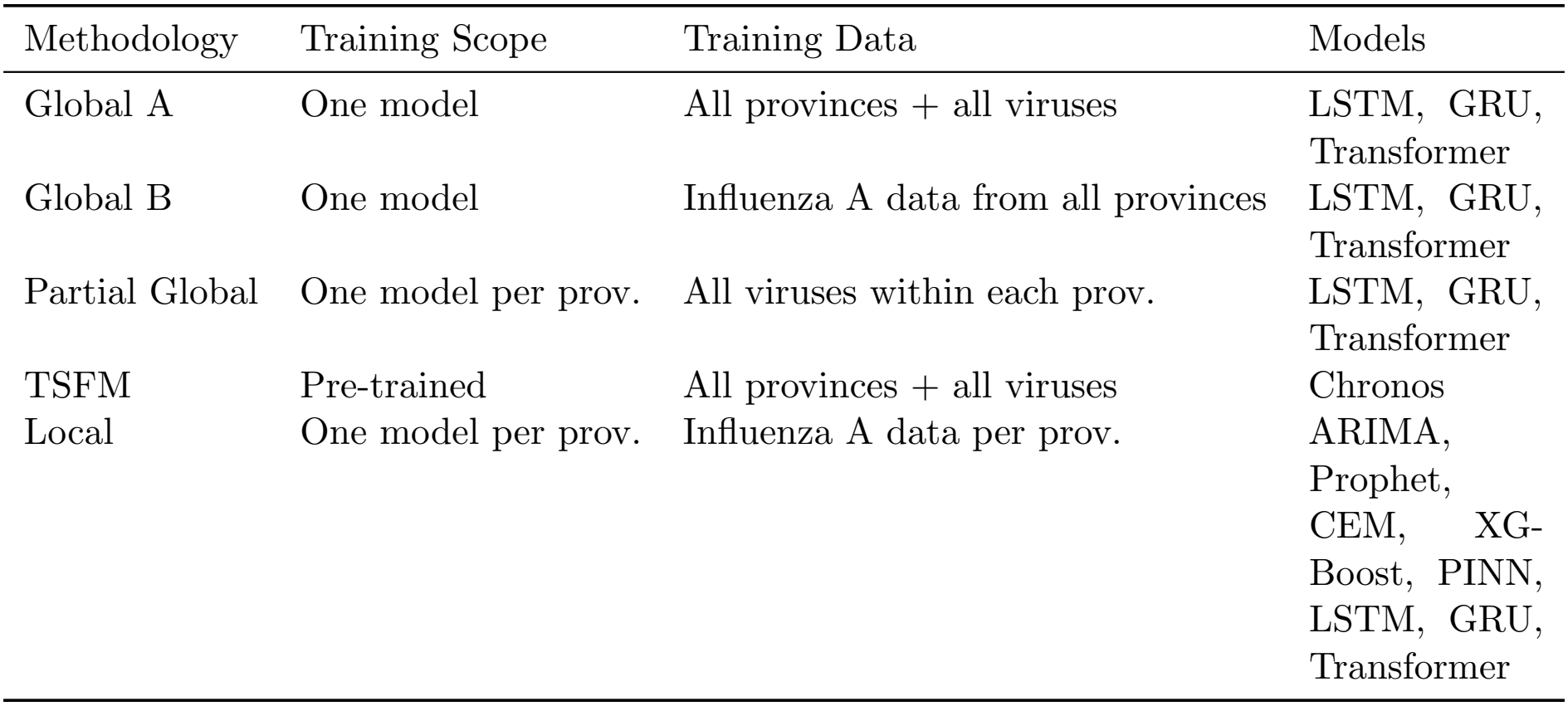
Summary of forecasting paradigms considered in this study.

Through these paradigms, we systematically assess whether cross-province and cross-virus data sharing improves downstream forecasting accuracy. We trained statistical, ML, PINN, and CEM models locally, with a separate model fitted to each province for influenza A only. We trained DL models under both local and global settings, allowing us to compare province-specific learning with joint learning across provinces. TSFMs are global models by nature, corresponding to Global A in our setting, because they are pre-trained on datasets from different fields. The details are provided in Table 2.

Although the global models are trained on data from multiple diseases and/or provinces, we tune hyperparameters and evaluate models exclusively for the target disease, influenza A. During forecasting, the model uses only influenza A positivity-rate data, with forecasts generated independently for each province.

### 3.1 Data preparation and splitting

For each province, we independently normalize the influenza A time series using Min-Max scaling and transform it into a supervised learning dataset through lag embedding. The lagged observations serve as predictor variables, while the subsequent observation serves as the target variable. We then train a separate local model for each province using only the corresponding influenza A lag-embedded matrix.

For global modeling, individual time series are scaled (Min-Max or standardized), transformed into lagged feature matrices, and vertically stacked into a single supervised regression dataset [73].

We train the models on the training set, tune their hyperparameters on the validation set, and evaluate their forecasting performance on the test set. The dataset, comprising data from the 2007/2008 season through the 2025/2026 season, is evaluated using a rolling-window procedure. The forecasting period begins on October 11, 2025, and extends through March 28, 2026. Thus, October 4, 2025, is used as the as-of date to ensure that no observations from the forecasting period are used for model training or hyperparameter tuning.

For each as-of date, only observations available before and on that date are used to construct the training and validation sets. The initial training period ends on October 2, 2024, and the training end date is subsequently advanced by one week at each iteration. Consequently, the training period expands progressively throughout the evaluation period. At each iteration, observations from dates after the training end date through and including the as-of date are used as the validation set. Forecast performance is evaluated on observations in the test period, from October 11, 2025, through March 28, 2026. This is applied to all models.

### 3.2 Model training

All models considered in this study were trained and evaluated to generate deterministic *point forecasts* across the specified horizons. We use a recursive strategy for ML and DL models.

- XGBoost is evaluated for point forecasting. We use a standard regression objective and optimize the model parameters directly for continuous target regression. The XGBoost hyperparameters are optimized using Optuna [74].
- We evaluate LSTM, GRU, and the Transformer architecture for point forecasting using dense output layers to produce continuous predictions. Furthermore, we fix the number of training epochs at 1,000 and use the Adam optimizer [75] for training.
- We estimate the ARIMA model parameters (*p, d, q*) from the historical series to capture underlying temporal dependencies and seasonality, producing deterministic point forecasts for each forecast horizon. We select *p, d, q* that produced the lowest Akaike Information Criterion (AIC).
- For Prophet, we generate point forecasts directly from the fitted statistical model using its model-specific forecasting mechanism. The underlying additive model that decomposes a time series into trend, seasonal, and holiday components, producing deterministic point predictions across each forecast horizon.
- We evaluate Chronos for point forecasting under three settings: (1) zero-shot forecasting, (2) LoRA (Low-Rank Adaptation) fine-tuning, which updates a subset of model parameters, and (3) full fine-tuning, which updates all model weights. For each setting, we derive point forecasts directly from the model’s predicted autoregressive output distribution at each forecast horizon.

Table 3 provides the full hyperparameter search space and tuning configurations for each model.

**Table 3:** Hyperparameter search spaces for deep learning, machine learning, Prophet, and Chronos models.

| Model | Hyperparameter | Search space |
|---|---|---|
| LSTM | Hidden size | {32, 64, 128} |
|  | Number of layers | {1, 2, 3} |
|  | Dropout | [0, 0.3] |
|  | Learning rate | [10<sup>−4</sup>, 3 × 10<sup>−3</sup>] (log-uniform) |
|  | Batch size | {16, 32, 64} |
| GRU | Hidden size | {32, 64, 128} |
|  | Number of layers | {1, 2, 3} |
|  | Dropout | [0, 0.3] |
|  | Learning rate | [10<sup>−4</sup>, 3 × 10<sup>−3</sup>] (log-uniform) |
|  | Batch size | {16, 32, 64} |
| Transformer | Model dimension (*d*<sub>model</sub>) | {32, 64, 128} |
|  | Attention heads (*n*<sub>head</sub>) | {2, 4, 8} |
|  | Number of layers | {2, 3, 4} |
|  | Feed-forward dimension | {8, 16, 32, 64, 128, 256} |
|  | Dropout | [0, 0.3] |
|  | Learning rate | [10<sup>−4</sup>, 3 × 10<sup>−3</sup>] (log-uniform) |
|  | Batch size | {16, 32, 64} |
| XGBoost | Number of estimators (*n*<sub>estimators</sub>) | {100, 200, 400} |
|  | Learning rate | [0.01, 0.2] (log-uniform) |
|  | Maximum tree depth (max_depth) | {3, . . . , 10} |
|  | Subsample ratio | [0.6, 1.0] |
|  | Column subsampling ratio | [0.6, 1.0] |
|  | Minimum child weight | {1, . . . , 10} |
|  | Minimum split loss (*γ*) | [0, 5] |
|  | L2 regularization (*λ*) | [10<sup>−3</sup>, 10] (log-uniform) |
| Prophet | Growth | {linear} |
|  | Number of changepoints | {50, 55, 60, . . . , 100} |
|  | Changepoint range | [0.99, 1.0] |
|  | Seasonality mode | {multiplicative} |
|  | Seasonality prior scale | [0.1, 15.0] |
|  | Changepoint prior scale | [0.001, 2.0] (log-uniform) |
|  | Holidays prior scale | [0.1, 10.0] |
|  | Uncertainty samples | {500, 1000, 1500} |
|  | Yearly seasonality | {True} |
|  | Weekly seasonality | {False} |
|  | Yearly Fourier order | {8, 10, 12, 15} |
|  | Weekly Fourier order | {3, 5, 7} |
| Chronos | Learning rate | {10<sup>−4</sup>, 10<sup>−3</sup>, 10<sup>−2</sup>} |
|  | Batch size | {32, 64} |
|  | Number of steps | {200, 500, 1000} |
|  | Context length (*p*) | {2, 4, 8, 16} |

**Table 4:**
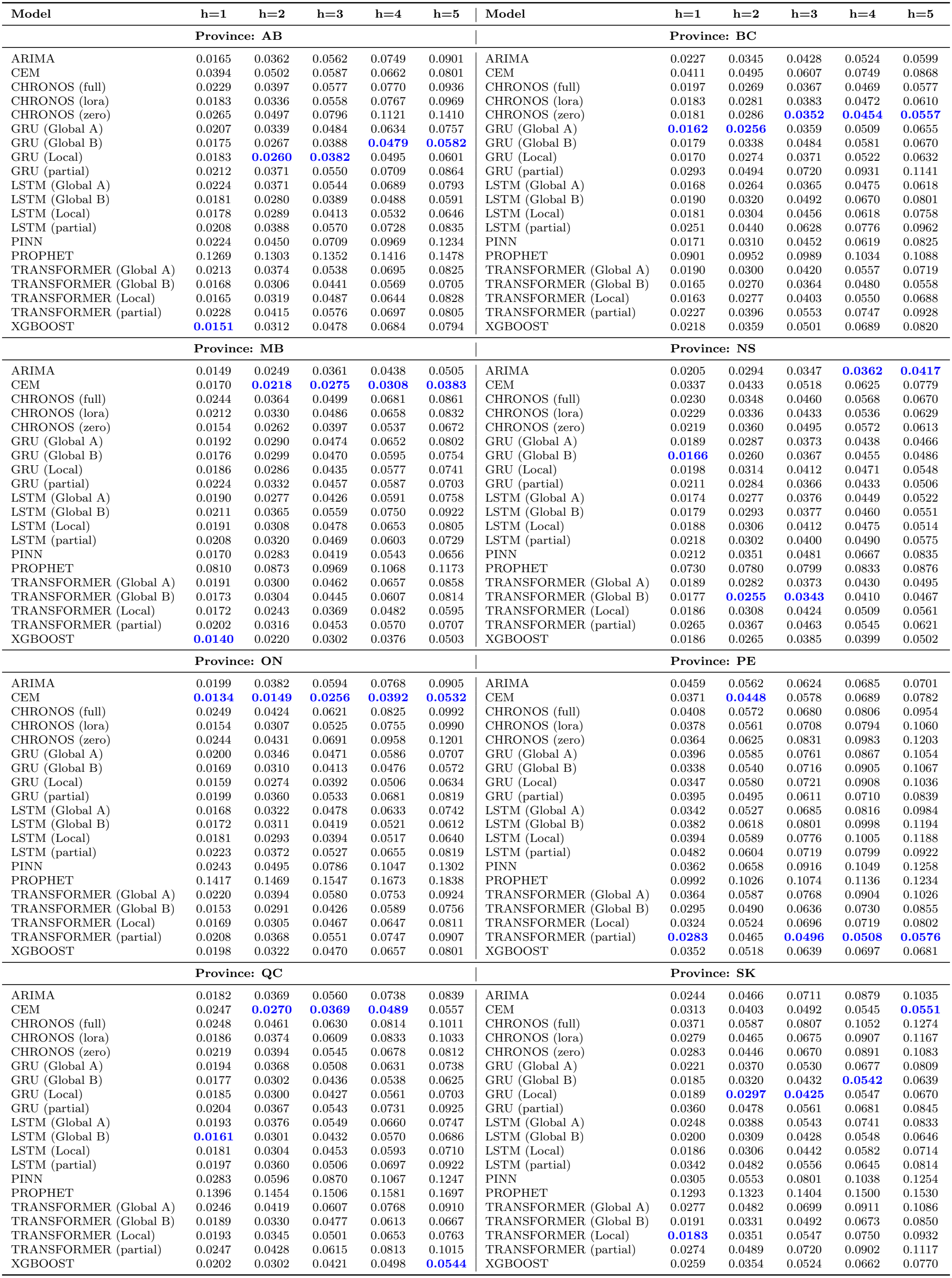
MAE Summary across Horizons (*h* = 1 to *h* = 5) by Province (Lowest MAE highlighted)

**Table 5:** Machine learning types, problems, and representative models.

| Learning Type | Problem | Model |
|---|---|---|
| Supervised | Classification | Logistic Regression, k-NN, Decision Trees, SVM, Neural Networks (NNs) |
|  | Regression | Linear Regression, Polynomial Regression, SVR, NNs, Decision Trees |
| Unsupervised | Clustering | K-Means, Hierarchical Clustering, DBSCAN |
|  | Dimensionality Reduction | PCA, t-SNE, UMAP, Autoencoders |
|  | Anomaly Detection | Isolation Forest, DBSCAN, One-Class SVM, Autoencoders |
| Reinforcement | Policy Optimization | Q-Learning, Deep Deterministic Policy Gradient (DDPG) |

### 3.3 Hyperparameter tuning

Due to computational constraints, we tuned hyperparameters for the machine learning and deep learning models only once at the initial as-of date using Optuna across 100 trials to minimize validation loss. The optimal parameters were then fixed for all subsequent rolling forecasts. Conversely, because the statistical models were much less computationally expensive, we retuned their hyperparameters at every as-of date.

The model will be retrained at each subsequent as-of date with the updated training data and the same hyperparameters chosen at the first as-of date. The forecast will be obtained from the trained model for the following five steps, where the test set provides the actual values. We apply this forecasting procedure to all as-of dates to mimic a real-life scenario.

The details presented in this section apply to the AI and statistical models, while the PINN and CEM are described separately in their respective self-contained sections.

### 3.3 Compartmental epidemiological model

In this study, we use a relatively simple compartmental model using a discrete time renewal equation implementation (which is mathematically equivalent to the more traditional implementation using ODEs [7]). This is a model that has been used internally at the Public Health Agency of Canada during the past 5 years to forecast seasonal acute respiratory infections in Canada. The observed incidence *i*_*t*_ on week *t* is defined with the following equations:

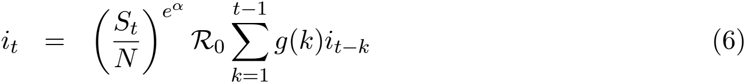

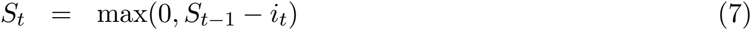

where *S*_*t*_ is the number of individuals still susceptible to infection at time *t, N* is the total population size, *ℛ*_0_ is the basic reproduction number, *g* is the intrinsic generation interval distribution [76] and *α* is a parameter representing the heterogeneity on contacts (*α* = 0 for homogeneous mixing of the population). The model parameters *R*_0_ and *α*, as well as the initial condition *i*_0_ are fitted to the observed incidence of the *currently emerging* season. The model is used for each selected provinces, independently. For seasonal influenza, the distribution *g* was set as a gamma distribution with mean 3 days and variance 1 day. The population size *N* was retrieved from the Statistics Canada table associated with the province. For calibration, the CEM uses only a limited amount of past data from the current 2026 season; data from previous seasons are not used. The calibration algorithm used is an Approximate Bayesian Computation with 10,000 iterations that selects the associated parameters of the 100 trajectories closest to the fitting data set (top 1%).

### 3.4 PINN and training details

To incorporate epidemiological domain knowledge into the neural network without lag embedding, we constrain the learned dynamics using the SIR model. As illustrated in Figure 6, the network NN(*t*; *θ*) maps time input *t ∈* [0, 1] to the state variables [*S*(*t*), *I*(*t*), *R*(*t*)]^*⊤*^. Target incidence data are min-max normalized to [0, 1] for optimization stability and inverse-transformed for post-training forecasting. To enforce non-negativity, parameter estimates 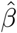 and 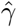 are parameterized logarithmically (initialized at 0.80 and 0.10, respectively) and jointly optimized alongside the network weights over 12,000 full-batch epochs.

**Figure 6.**
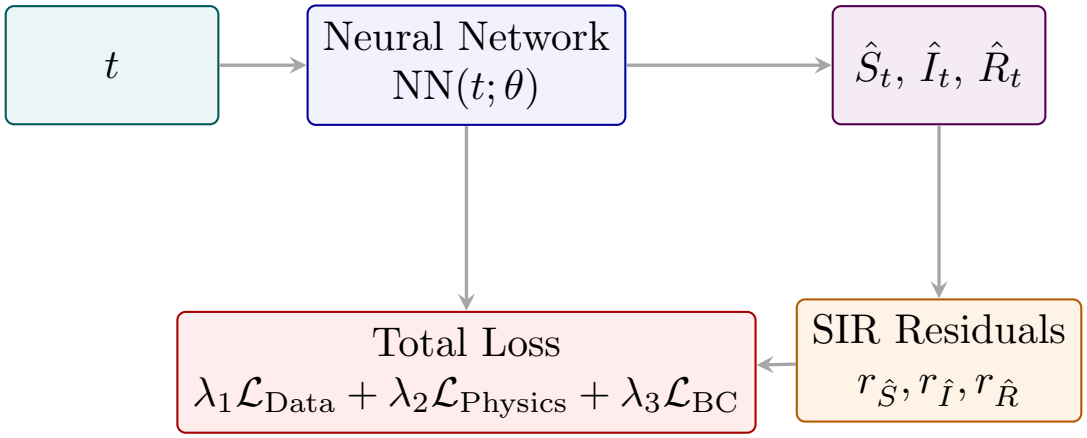
Architecture of the SIR PINN used in this study.

The PINN is trained by minimizing the weighted composite loss function

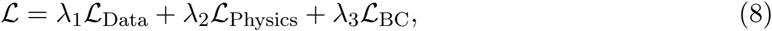

where each loss component is defined as follows:

#### Data Loss

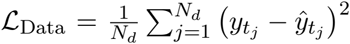, where 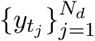 denotes the observed influenza A incidence data and 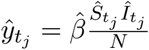 is the predicted incidence at time *t*_*j*_.

#### Physics Loss

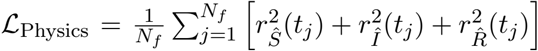, evaluated over *N*_*f*_ collocation points with physics residuals:

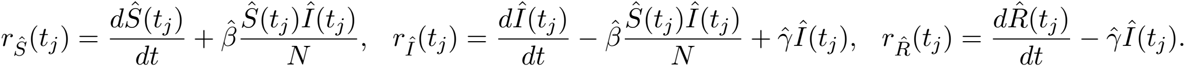

#### Boundary/Conservation Loss

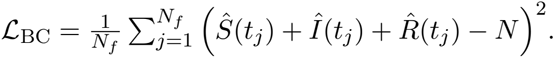.

Physics constraints are enforced via *N*_*f*_ = 3000 collocation points densely sampled across *t ∈* [0, 1], enabling fine-grained residual evaluation independent of sparse data observations. The objective loss weights are configured to *λ*_1_ = 1.0 (data), *λ*_2_ = 10^*−*4^ (physics), and *λ*_3_ = 1.0 (conservation). The smaller *λ*_2_ weight ensures that physics residuals guide learning without overwhelming data fidelity or total population constraints.

### 3.5 Forecast evaluation metrics

Our forecasting target is the influenza A positivity rates at horizons *h ∈ {*1, 2, 3, 4, 5*}* weeks ahead. Here, *t*^*⋆*^ denotes the as-of date (week). We evaluate point forecasts using the Mean Absolute Error (MAE):

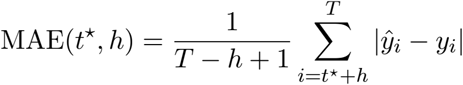

where *ŷ*_*i*_ denotes the predicted point forecast at time index *i* and *y*_*i*_ represents the corresponding observed value. Additionally, *T − h* + 1 is the number of forecasts or observations being evaluated for horizon *h*.

## 4 Results and Discussion

We report MAE for CEM, ARIMA, Prophet, and XGBoost in the local paradigm; GRU, LSTM, and Transformer in the local, global (A and B), and partial paradigms; and Chronos for zeroshot forecasting and two fine-tuning strategies: LoRA and full fine-tuning.

### 4.1 Experimental protocol

All experiments were implemented in Python (version 3.12.13). The computational experiments were performed on an internal high-performance computing cluster provided by the Public Health Agency of Canada. The computational resources used included Intel(R) Xeon(R) Gold 6130 CPU @ 2.90GHz, 32 CPU cores, and 128GB of memory. No GPU was used.

### 4.2 Results

This section presents the forecasting results obtained from the models considered in this study. We evaluate their performance in terms of point-forecast accuracy forecast performance across the eight Canadian provinces and the considered forecasting horizons. Specifically, we report the MAE.

For the window size-based models, we select the window size that yields the lowest MAE for each horizon. Then, we report in Figure 7 the sum of MAEs across horizons for each province, where the CEM model is chosen for benchmarking, as it has been the standard method in epidemic forecasting for many years.

**Figure 7.**
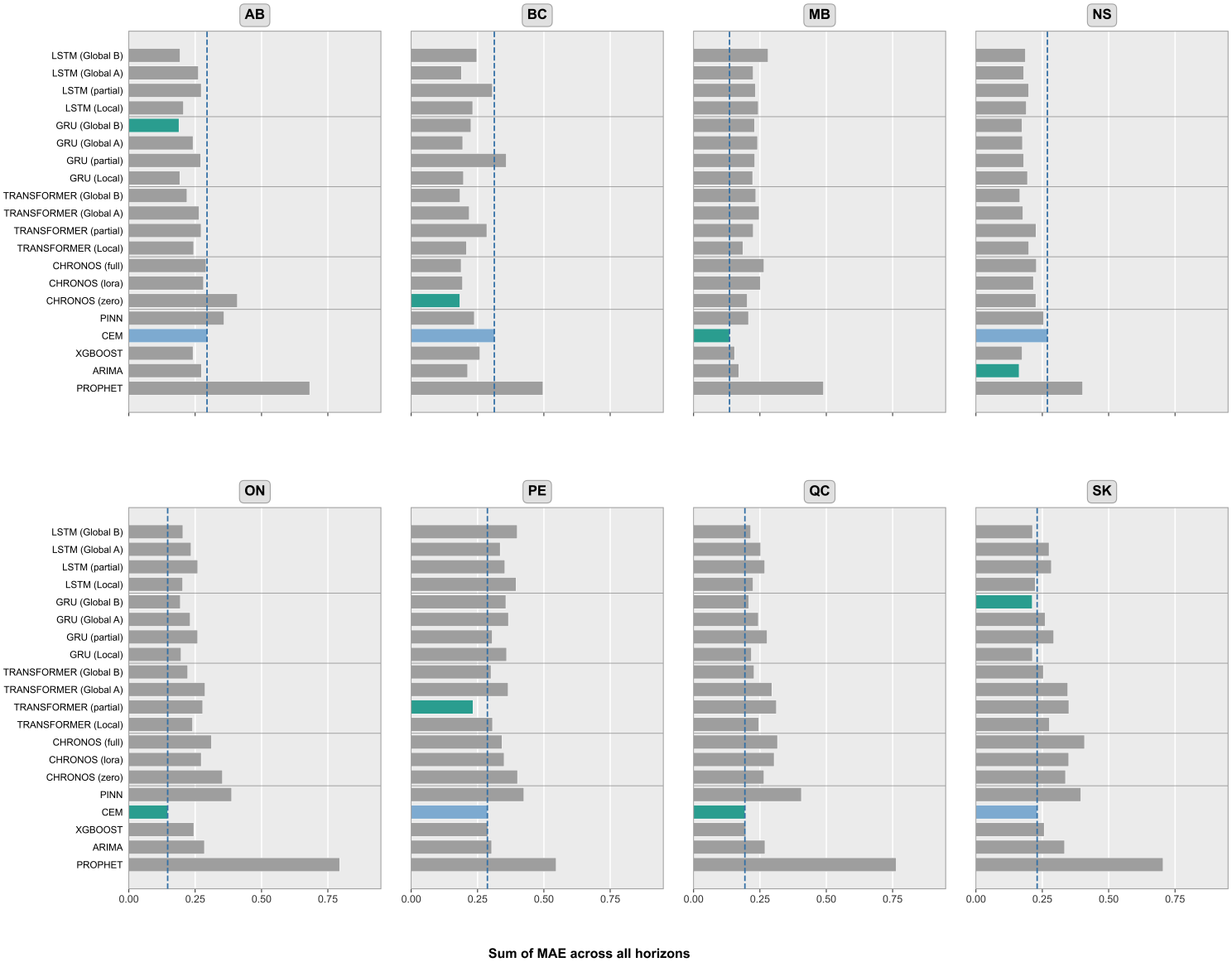
Mean absolute errors of the forecasted influenza A positivity for each model, by province. For each province, the best model is in green. The best model is in green. The blue vertical line extends to the CEM MAE to facilitate comparison.

**Figure 8.**
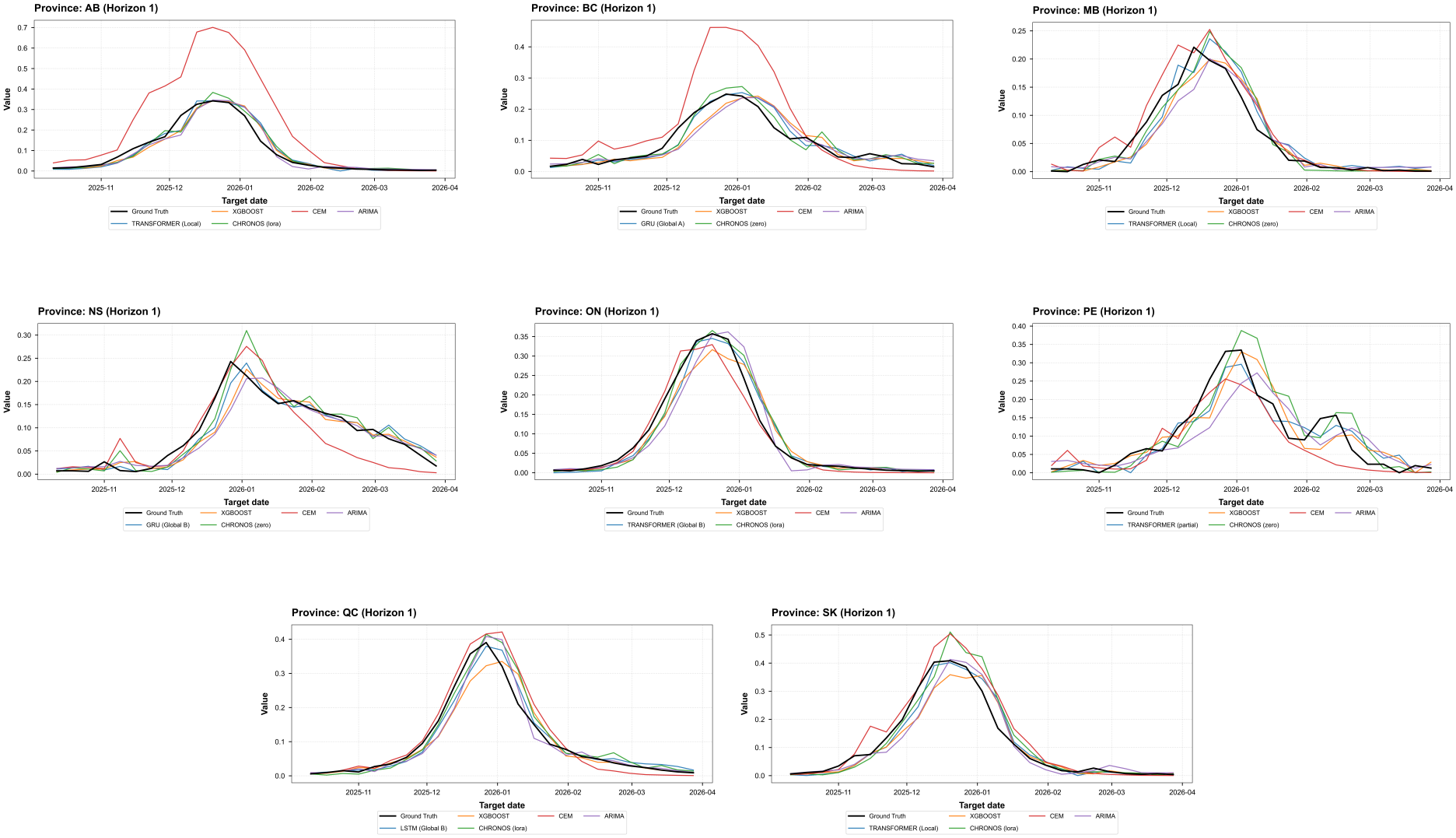
Forecast vs. Ground Truth: Comparison of the top-performing model per category across provinces.

**Figure 9.**
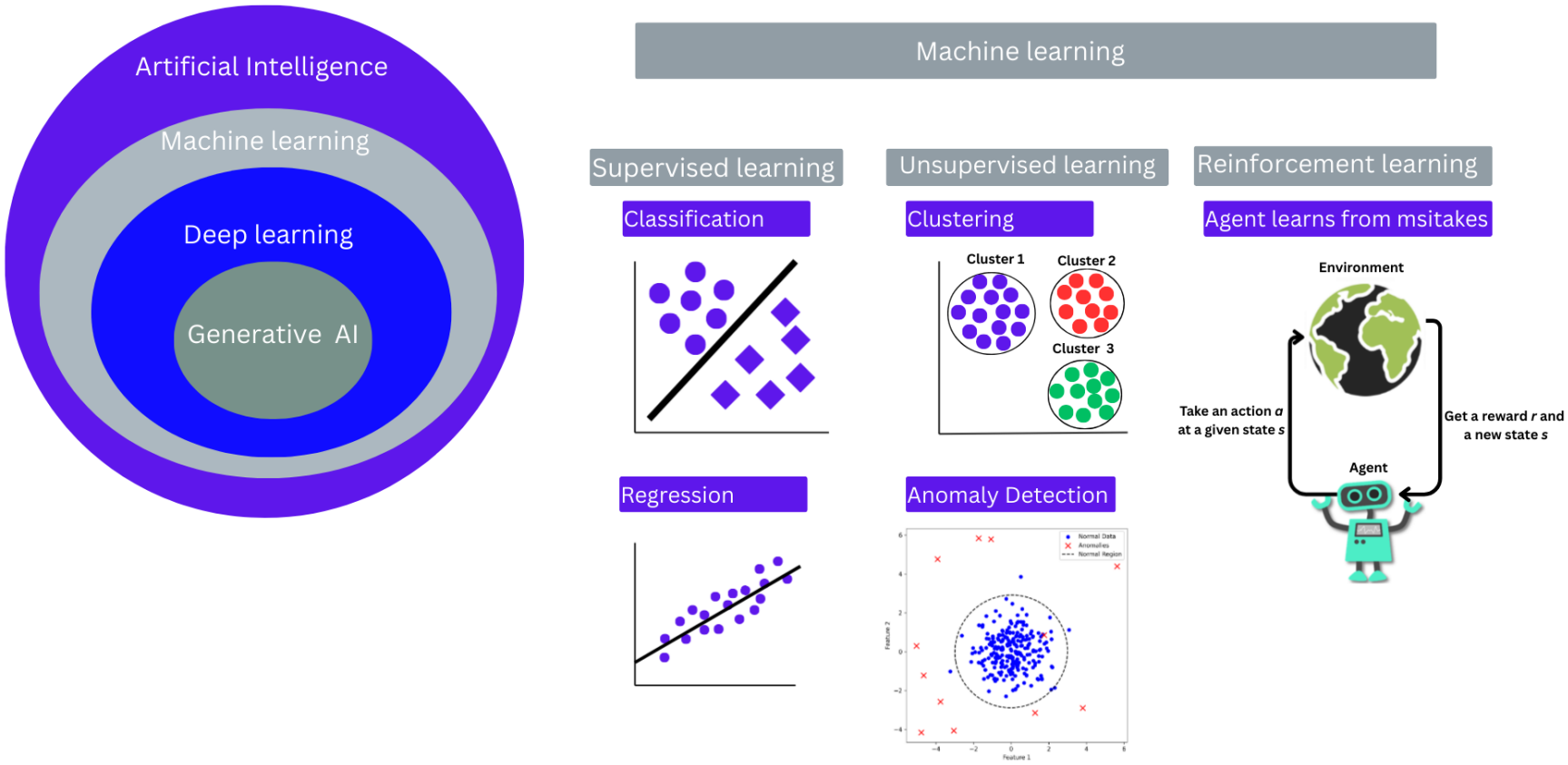
Artificial Intelligence (AI), Machine Learning (ML), Deep Learning (DL), and Generative AI: summary of learning types and associated problems. Supervised learning includes classification and regression problems; unsupervised learning includes clustering, dimensionality reduction, and anomaly detection; reinforcement learning involves learning through interaction with an environment and feedback.

**Figure 10.**
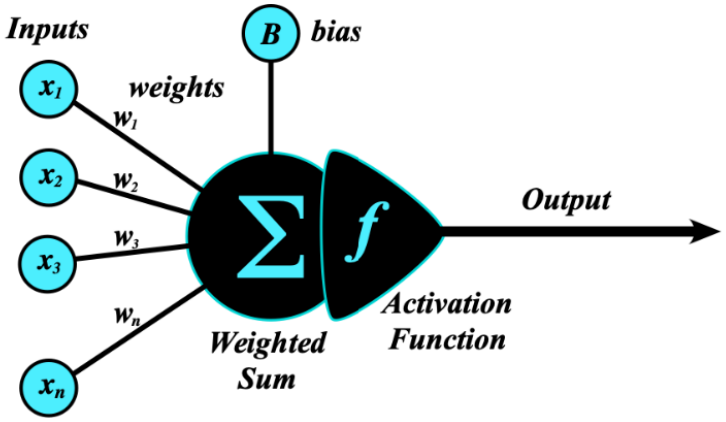
Schematic of a simple artificial neural network (ANN), consisting of an input layer and a single output neuron.

**Figure 11.**
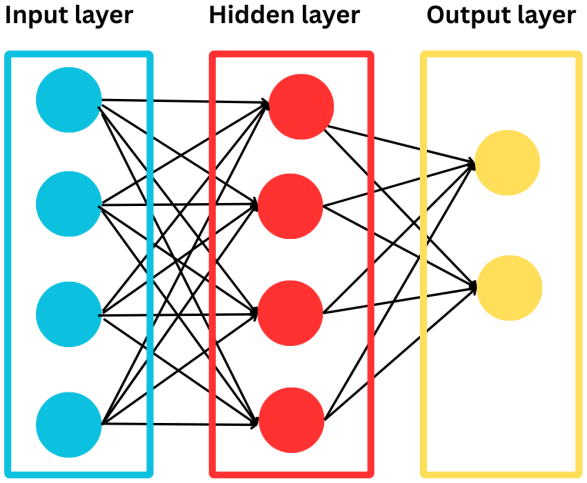
Schematic of a typical MLP with input, hidden, and output layers. Hidden layers learn intermediate feature representations.

**Figure 12.**
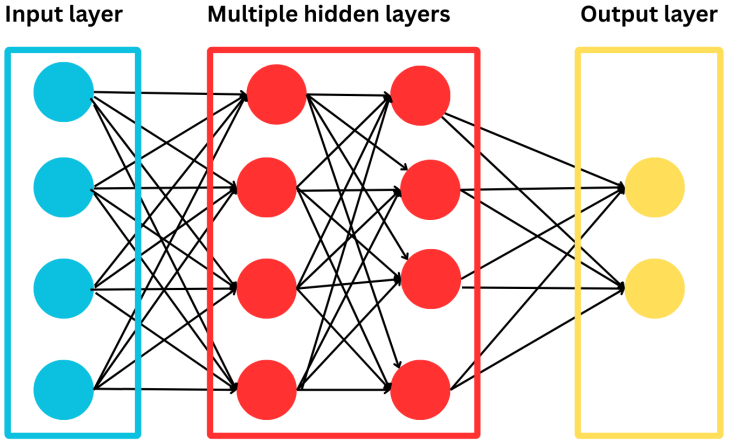
Schematic of a typical deep neural network architecture, showing an input layer, two hidden layers, and an output layer. Hidden layers learn intermediate feature representations from the input data.

The CEM outperforms all other selected models in Ontario, Quebec, and Manitoba (relatively high-population provinces), showing that the mechanistic constraints prevent it from overfitting to noise during the season. Regarding deep learning models, cross-provincial and cross-virus data pooling substantially improves performance; specifically, globally and partially pooled models, such as GRU (Global B), which outperforms all other selected models in Al-berta and Saskatchewan, and Transformer (partial) in Prince Edward Island, demonstrate that capturing shared cross-provincial and multi-virus dynamics improves upon purely deep learning models trained only on the target virus (FLUA). Remarkably, the Chronos (zero-shot forecasting) model is the best in British Columbia, and the ARIMA statistical model in Nova Scotia. In contrast, Prophet consistently underperforms and has the highest overall MAE. Similarly, PINN models underperform compared to both classical CEM and top-performing deep learning models.

### 4.3 Discussion

The exceptional growth of machine learning models underpinning “artificial intelligence” has driven a revolution in what science thought possible: large language models, protein-folding algorithms, and computer coding agents are just a few examples. Hence, it is natural to apply these innovations in epidemiology, particularly to support efforts to lessen the impact of seasonal acute respiratory infections, which significantly strain health systems. Hence, our study aimed to answer a simple question: as of 2026, can “AI” models forecast relatively standard seasonal epidemics, and can they outperform canonical compartmental epidemiological models (CEMs)? As a concrete example, we chose to forecast influenza A in selected Canadian provinces.

To do this, we reviewed the literature and selected machine learning models that already yielded promising results in other studies. After the literature review, we included one tree-based ensemble model (XGBoost), three deep learning models (LSTM, GRU, and Transformer), a physics-informed neural network (PINN) model, and one time-series foundation model (Chronos). To compare these AI models with more traditional approaches, we chose an ARIMA model, a simple yet popular statistical approach; a more sophisticated statistical model, Prophet; and a SIR-like compartmental mechanistic model (CEM) used internally within PHAC to forecast seasonal respiratory infections over the past 5 years.

Short-term forecast accuracy for AI models was comparable to, and in select provinces marginally better than, traditional approaches; similarly, accuracy declined as the forecast horizon increased. However, when accuracy was better, it was only marginally so, which ultimately does not really compensate for their limitations presented in subsection 2.10. As a result, the expectation for the level of forecast accuracy should be much higher for AI models than for mechanistic models; in other words, to be practically useful, AI models should be much more accurate to compensate for their limitations (the main one being their intractability). Additionally, we found that documentation for implementing AI models for non-expert users was minimal, if not lacking, for most AI models, consequently hampering their use by public health practitioners. Furthermore, we note that the expertise required to run most AI models is currently relatively high, hampering adoption in most public health settings because they generally lack highly technical staff.

An important finding from our study is that while AI models could reach a similar or better accuracy than the CEM in half of the provinces (e.g., GRU, Transformer, and Chronos), the improvement was only marginal and, from a practical viewpoint, did not compensate for the opaqueness of AI models. Indeed, public health practitioners need to understand the basic mechanisms behind a given forecast to explain the underlying epidemiology and, when forecasts are pessimistic, provide potential countermeasures to decision-makers. Given their mechanistic nature, CEMs can disentangle all epidemiological aspects of a forecast, whereas AI models are largely inscrutable.

Although PINN models did not perform best in our study, they may be a promising approach in some cases. This lower performance may stem from the simple SIR model it uses, or from the difficulty of tuning the loss weights. Future work should thoroughly analyze how these loss terms affect PINN accuracy.

While TSFMs had a promising performance, Chronos achieved the best performance in British Columbia under zero-shot settings, demonstrating its utility without task-specific fine-tuning. Nevertheless, we do not see Chronos systematically outperforming the best-performing models across all jurisdictions. Future work should explore more TSFMs based on architectures (including transformer variants, i.e., encoder-decoder, encoder-only, and decoder-only, non-transformer models, and diffusion models), learning strategies (fully supervised vs. self-supervised learning), and input representations (e.g., point-based vs. patch-based tokenization), as these factors may affect performance. Synthesizing those factors will establish benchmark practices for epidemic forecasting.

In this work, we explored cross-domain generalization by pooling time-series data from multiple provinces and viruses (Influenza B, RSV, and COVID-19). This data pooling significantly improved deep learning model performance, enabling globally and partially pooled architectures (such as GRU and Transformer) to outperform localized variants and achieve top ranks in multiple provinces. Expanding cross-domain data pooling remains a critical avenue for future research in epidemic forecasting.

Despite the advantages of CEMs in providing insights into disease progression and their inherent interpretability, several limitations remain. First, CEMs often rely on simplifying assumptions, such as homogeneous population mixing (even when the population is stratified), simplified contact patterns, or simplified pathogen evolution. Second, model misspecification and partial observability may occur when the chosen CEM does not fully reflect real-world dynamics or when some compartments are not directly observable. Third, epidemiological data are often sparse and uncertain because of under-reporting, testing limitations, and reporting delays, which complicates estimation of key model parameters. These parameters are typically inferred from historical data that may change over time due to evolving public health policies, pathogen evolution, behavior change, etc.

Our study focused narrowly on the 2025/2026 influenza A season in selected Canadian provinces. Hence, our results may not be generalizable to other settings or viruses. Although we chose the Canadian setting out of interest, seasonal respiratory virus epidemic waves in Canada are similar to those in other temperate western countries. Because AI models are data-driven, we expect similar findings if we applied our study to other similar geographical locations. Forecasting influenza A, rather than other seasonal respiratory viruses, was chosen as we thought this virus provided a good balance of complexity: on the one hand, access to historical data of about 15 past seasons likely facilitated the fitting and training of the forecasting models; on the other hand, there is a marked heterogeneity between influenza A seasons (e.g., season start and peak timing, growth rate, etc.), likely caused mainly by the rapid evolution of this virus.

We attempted to leverage the AI models well. However, our use may not have been optimal and may have impacted forecasting performance. In future work should explore other supervised methods for generating samples, such as direct, multi-output, or hybrid models.

This study emphasized a practical approach: using AI models in a typical public health setting by professionals with a fair understanding of machine-learning theory and implementation, but who are not experts in this field.

## 5 Conclusion

Evaluating the 2025/2026 influenza A season across Canadian provinces shows that short-term forecast accuracy for AI models (such as GRU, Transformer, and Chronos) was comparable to or better than traditional models, and each model’s accuracy declined as the forecast horizon increased. The empirical findings yield key insights for AI deployment: cross-provincial and cross-virus data pooling help AI models to mitigate overfitting on small local sample sizes; AI models are slightly better than CEM, and zero-shot TSFMs like Chronos are also promising. Conversely, models like Prophet or PINNs performed poorly. An important finding from our study is that while AI models could reach similar or better accuracy than the CEM, the improvement was only marginal and, from a practical viewpoint, did not compensate for AI models’ limitations. Because public health practitioners require transparent, epidemiologically grounded forecasts to explain disease dynamics and justify interventions to decision-makers, mechanistic CEMs remain the most practical and reliable primary forecasting tool.

As AI models improve and become more widely used, they could play an important role in epidemic forecasting. Further directions for investigation are numerous. One approach is to develop hybrid methods that combine AI models with traditional models (statistical models and CEMs) to exploit their complementary strengths. Alternatively, AI models can enhance CEMs by inferring their parameters from the observed data. Such approaches could combine AI’s predictive power with the interpretability and epidemiological structure of mechanistic models. Another route is to use ensembling forecasts from different models to improve accuracy of forecasting. Combining forecasts can reduce model-specific errors and improve robustness across provinces, forecast horizons, and epidemic seasons.

## A AI-based approaches

Artificial Intelligence (AI) refers to the capability of computer systems to perform tasks that typically require human intelligence, such as reasoning, visual perception, decision-making, problem-solving, and learning, among others. Machine learning (ML) is a subfield of AI that enables a system to learn a function *f* : *X →Y* from historical data 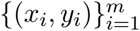 and improve its performance without being explicitly programmed [24].

AI and ML have been used in epidemics for various tasks, including outbreak detection, Infectious disease surveillance, Public health decision-making, Scenario modeling, transmission pattern prediction, and epidemic forecasting. In this paper, we focus on epidemic (infectious disease) forecasting task. Real-time epidemic forecasting enables the prediction of geographic disease spread, case counts (incidence), hospitalizations, and deaths, thereby supporting timely and evidence-based public health interventions [77].

AI-based approaches for epidemiology represent a powerful tool for improving disease fore-casting, particularly ML and deep learning (DL) methods. Unlike compartmental epidemiological models (CEMs), which rely on predefined ordinary differential equations (ODEs), AI-based approaches can learn patterns directly from historical incidence data. This capability enables them to capture complex relationships and generate forecasts of disease incidence, hospitalizations, and deaths.

### A.1 Machine learning

Machine learning (ML) is a subfield of AI that enables a system to learn a function *f* : *X →Y* from historical data 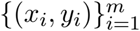 and improve its performance without being explicitly programmed [24]. In *supervised learning*, each input *x*_*i*_ *∈ X* has a corresponding target *y*_*i*_ *∈ Y* (*y*_*i*_ = *f*^*\**^(*x*_*i*_), *f*^*\**^ is a function from an expert), and a learning algorithm learns a mapping *f* that predicts the output for new inputs; common tasks include classification (predicting discrete target, e.g., whether a disease is present in a region) and regression (predicting continuous values, e.g., disease incidence over time). A learning algorithm (e.g., Gradient descent) learns the best 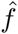 given the historical data. The resulting model captures the relationship between the input and output, and is able to predict a target of an input. In this work, we focus on regression, as forecasting aims to estimate a continuous future values. In *unsupervised learning*, only inputs *{x*_*i*_*}* are given, and the model discovers patterns or structures *f* : *X →Z*, such as clustering similar data points, detecting anomalies, or reducing the dimensionality of complex datasets while preserving key information. In *reinforcement learning*, an agent interacts with an environment in state *s*_*t*_, takes actions *a*_*t*_, and receives rewards *r*_*t*_; the goal is to learn a policy *π* : *s 1↦ a* that maximizes cumulative reward, as in adaptive epidemic interventions or game-playing applications.

Other types of learning:

1. *semi-supervised learning* : combines supervised and unsupervised learning by using a small portion of labeled data together with a larger amount of unlabeled data, allowing the model to learn from both human-provided labels and unlabeled examples.
2. *self-supervised learning* : learns representations from unlabeled data by generating supervisory signals directly from the data itself, without requiring manually labeled examples.

Unsupervised learning has been applied to identify regions with similar patterns of disease activity [78], while reinforcement learning has been used to evaluate the effects of different intervention strategies over time. Supervised classification models have been widely employed for outbreak detection, whereas epidemic forecasting is treated as a regression problem. Commonly used supervised regression models in this domain include linear regression, support vector regression (SVR), decision trees, and neural networks, with ensemble approaches such as Random Forest and Extreme Gradient Boosting (XGBoost) frequently utilized for their predictive performance and robustness. Therefore, we consider a tabular dataset (e.g., a CSV file), where each input (row) has *n* characteristics or features, i.e., *x*_*i*_ *∈ X* = ℝ^*n*^. We briefly present some ML models.

#### Linear Regression

The linear regression model is 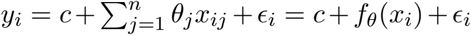, where *y*_*i*_ is the response for observation *i, x*_*i*_ = (*x*_*i*1_, …, *x*_*in*_) is the input vector for observation *i, x*_*ij*_ is the *j*-th input feature of *x*_*i*_, *θ*_*j*_ are the coefficients, *c* is the intercept, and *ϵ*_*i*_ is the error term. The goal is to find 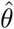 that minimizes the average loss, i.e.,

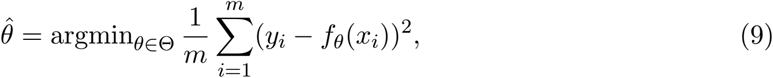

and this can be done by using an learning algorithm (e.g., Gradient descent or its variants). The function *f*_*θ*_ can be a neural network or any other machine learning regression models (except decision tree and ensemble methods, which are rule-based), and the goal remain the same.

Regularized versions of linear regression help prevent overfitting and improve model generalization by adding penalty terms to the loss function. Common methods include Ridge, Lasso, and Elastic Net.

To enable models to learn temporal dependencies from historical observations, the time series is transformed into supervised learning samples using lag embedding.

### A.2 Deep learning

Deep learning (DL) [79] is a subset of machine learning based on multi-layered neural networks capable of learning hierarchical representations of data. It has achieved strong performance across computer vision, natural language processing, speech recognition, and predictive analytics, with applications in healthcare, finance, and robotics.

#### Artificial Neural Network

An *Artificial Neural Network* (ANN) is a computational model inspired by biological neural systems, consisting of interconnected neurons organized in layers. These networks learn input-output mappings by adjusting weights and biases during training.

#### Neurons

Neurons are the basic computational units of an ANN. Each neuron computes a weighted sum of its inputs, adds a bias term, and applies a nonlinear activation function:

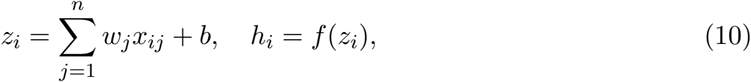

where *w*_*j*_ are weights, *b* is the bias, and *f* (*·*) is the activation function.

#### Activation functions

Activation functions introduce non-linearity, enabling the network to model complex relationships. Common examples include ReLU, sigmoid, and tanh. Without them, the network would behave as a linear model regardless of depth. The nonlinear activation function can be

1. Relu: *h* = *σ*(*z*) = max(0, *z*)
2. Tanh: 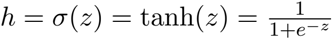
3. Sigmoid: 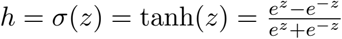

#### Layers

ANNs are organized into input, hidden, and output layers. The input layer receives data, the hidden layers extract intermediate representations, and the output layer produces predictions.

#### Feedforward neural network

A *feedforward neural network* (FNN) is an ANN where information flows in one direction from input to output without cycles, making it computationally efficient.

#### Multi-Layer Perceptron

A *Multi-Layer Perceptron (MLP)* is a feedforward neural network with at least one hidden layer. It uses nonlinear activation functions and is trained using backpropagation and gradient-based optimization. A single hidden-layer MLP is also called a shallow neural network.

#### Deep Neural Network

A *Deep Neural Network (DNN)* extends the MLP by using multiple hidden layers, enabling hierarchical feature learning where lower layers capture simple patterns and deeper layers encode abstract representations.

A DNN computes a composition of nonlinear transformations:

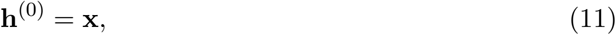

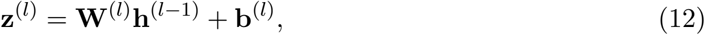

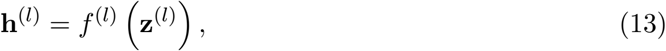

for *l* = 1, …, *L*, with final output:

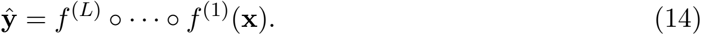

A standard Multi-Layer Perceptron (MLP) treats the input as a fixed-dimensional feature vector constructed from a window of past observations. For a lag order *p*, the input vector at time *t* is defined as

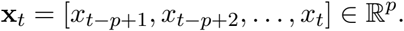

The MLP then learns a nonlinear mapping from the input space to the target value through a parametric function *f*_*θ*_. The prediction of the next observation is given by

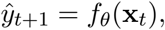

MLPs are not inherently designed to model temporal dependencies because they treat time series inputs as fixed feature vectors rather than ordered sequences (see Figure 13). For example, given a window **x**_*t*_ = [*x*_*t−*2_, *x*_*t−*1_, *x*_*t*_] to predict *ŷ*_*t*+1_, an MLP processes the lagged observations jointly as a static input without explicitly modeling their temporal order or retaining information beyond the input window. Moreover, the number of model parameters increases with the input dimension, which can increase computational cost and the risk of overfitting. These limitations make MLPs less suitable for capturing temporal dependencies in time series forecasting.

**Figure 13.**
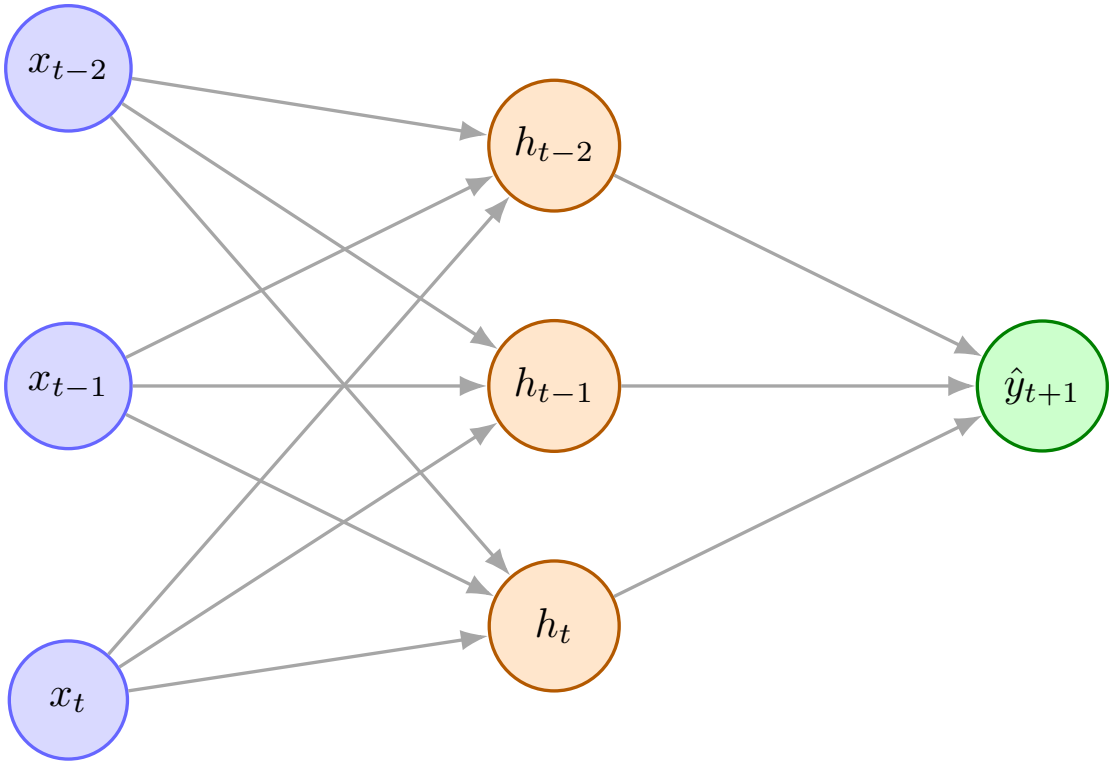
MLP for time series forecasting. The observations [*x*_*t−*2_, *x*_*t−*1_, *x*_*t*_] are treated as a static input vector to predict *ŷ*_*t*+1_.

**Figure 14.**
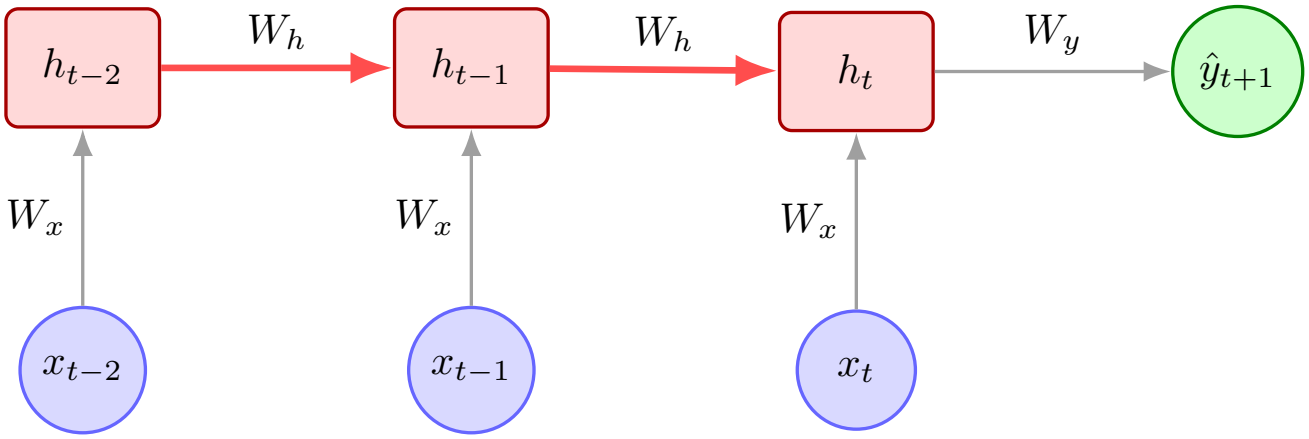
RNN for time series forecasting. Hidden states propagate temporal information recursively, enabling sequential dependency modeling. The weights *W*_*x*_, *W*_*h*_, and *W*_*y*_ are shared across time steps.

**Figure 15.**
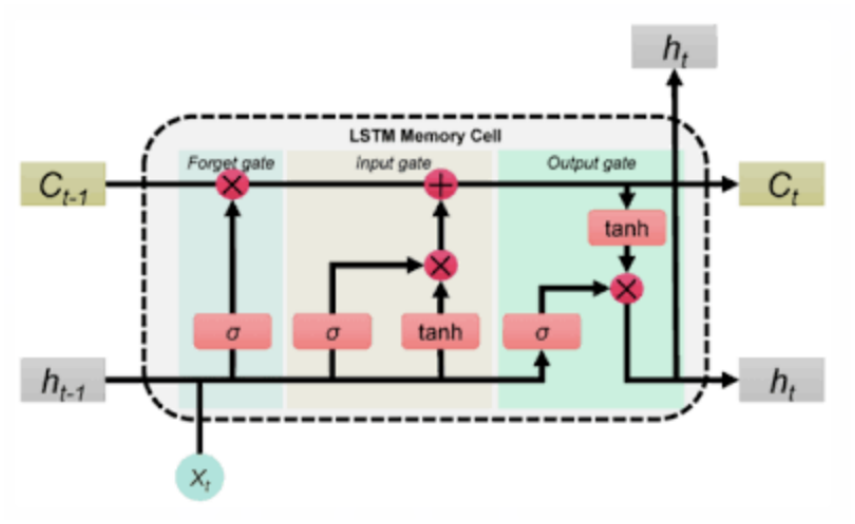
Schematic of an LSTM cell, illustrating the flow of information through the input, forget, and output gates, as well as the cell state and hidden state.

**Figure 16.**
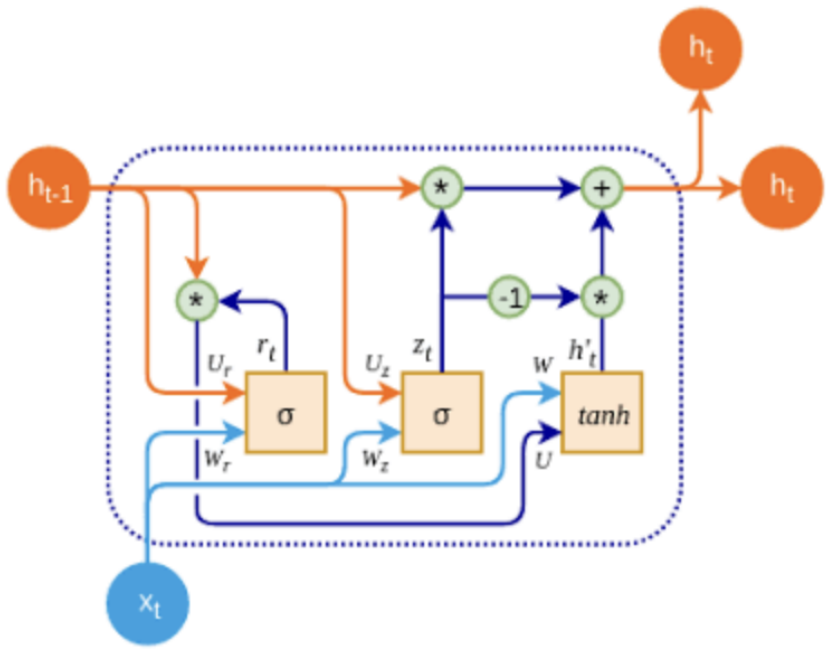
Schematic of a GRU cell, illustrating the flow of information through the update and reset gates, as well as the candidate activation and hidden state update.

**Figure 17.**
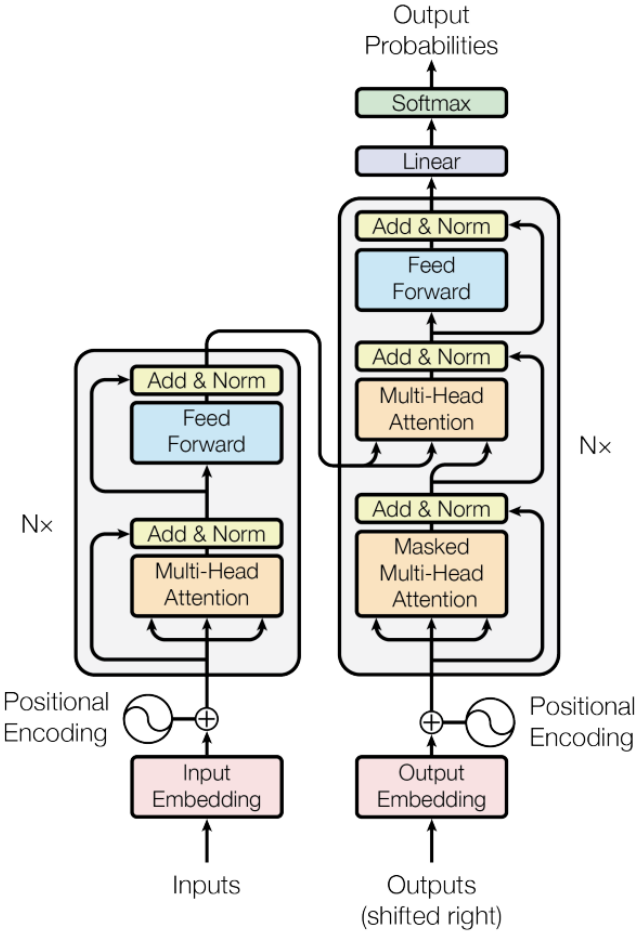
Schematic of the transformer architecture, illustrating the encoder-decoder structure and self-attention mechanism. The encoder processes the input sequence to generate contextualized representations, while the decoder uses these representations to produce the forecasted output.

#### Recurrent neural networks

To address the limitation of Feedforward Neural Networks (FNNs) or MLPs in handling sequential data, Recurrent Neural Networks (RNNs) introduce an explicit notion of *memory* through a hidden state. Let us consider the same example.

RNNs process the sequence step-by-step and maintain a hidden state *h*_*t*_ that acts as memory. It allows the model to capture temporal dependencies across time steps.

Starting with an initial state *h*_*t−*3_, the hidden states are updated recursively:

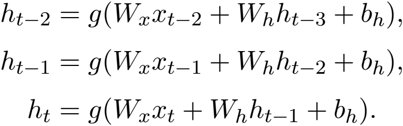

The prediction is then computed from the final hidden state:

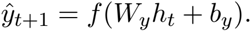

Here:

- *h*_*t−*2_ summarizes information from *x*_*t−*2_,
- *h*_*t−*1_ summarizes information from (*x*_*t−*2_, *x*_*t−*1_),
- *h*_*t*_ summarizes information from (*x*_*t−*2_, *x*_*t−*1_, *x*_*t*_).

Unlike DNNs, RNNs explicitly model temporal dependence through recursive hidden states, allowing past observations to influence future predictions through memory.

Although RNNs use a hidden state to model temporal dependencies, they can struggle to capture long-range relationships due to the vanishing gradient problem. As information is propagated through successive time steps, the influence of earlier observations diminishes, causing the hidden state to become increasingly dominated by recent observations. Consequently, RNNs may have difficulty capturing long-term and periodic patterns in long sequences, motivating the use of gated architectures such as LSTMs and GRUs, which regulate the flow and retention of information.

#### Long Short-Term Memory Networks

Long Short-Term Memory (LSTM) networks [37] are a specialized type of recurrent neural network (RNN) designed to mitigate the vanishing gradient problem that limits the ability of standard RNNs to learn long-range temporal dependencies. The name *Long Short-Term Memory* arises from the presence of two complementary memory representations within the architecture:

- the *cell state c*_*t*_, which acts as a *long-term memory* capable of preserving information across many time steps, and
- the *hidden state h*_*t*_, which acts as a *short-term memory* representing the current output and immediate contextual information.

Unlike conventional RNNs, LSTMs use gating mechanisms to regulate the flow of information into and out of the long-term memory state. These gates allow the network to selectively retain relevant information, discard irrelevant information, and incorporate new information during training.

The dynamics of an LSTM cell are governed by the following equations:

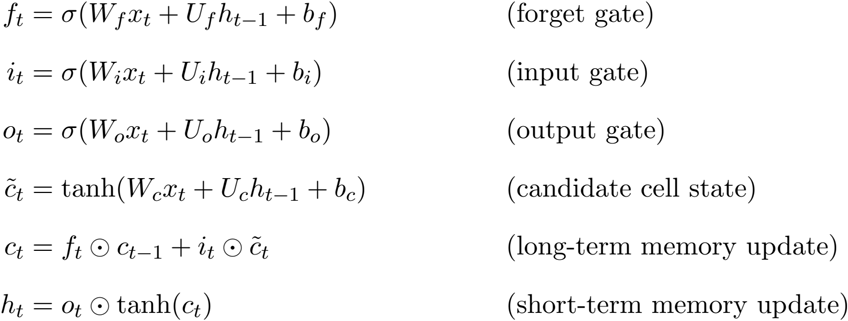

where:

- *x*_*t*_ denotes the input vector at time step *t*,
- *h*_*t−*1_ is the previous hidden state (short-term memory),
- *c*_*t−*1_ and *c*_*t*_ are the previous and updated cell states (long-term memory),
- *f*_*t*_, *i*_*t*_, and *o*_*t*_ represent the forget, input, and output gates,
- 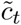 is the candidate cell state,
- *W*_*\**_ and *U*_*\**_ are trainable weight matrices,
- *b*_*\**_ are bias vectors,
- *σ*(*·*) denotes the sigmoid activation function,
- tanh(*·*) denotes the hyperbolic tangent activation function, and
- *⊙* denotes element-wise multiplication.

The long-term memory *c*_*t*_ enables the network to carry information over extended sequences with minimal modification, while the short-term memory *h*_*t*_ captures the information most relevant to the current time step. The forget gate determines which information from the previous long-term memory should be retained, the input gate regulates how new information is incorporated into the memory, and the output gate controls which parts of the long-term memory are exposed through the short-term memory. This architecture enables LSTMs to effectively model both short-range and long-range dependencies in sequential data.

#### Gated Recurrent Units

Gated Recurrent Units (GRUs) are a simplified variant of Long Short-Term Memory (LSTM) networks that also aim to address the vanishing gradient problem in standard Recurrent Neural Networks (RNNs). GRUs combine the forget and input gates of LSTMs into a single *update gate* and use a *reset gate* to control the influence of previous hidden states on the current candidate activation. This results in a more streamlined architecture with fewer parameters while still effectively modeling long-range dependencies in sequential data.

The GRU cell is defined by the following equations:

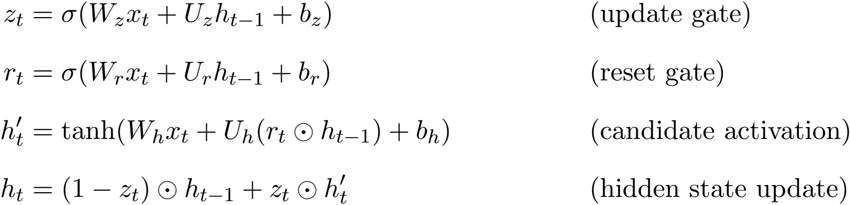

where:

- *x*_*t*_ is the input vector at time step *t*,
- *h*_*t−*1_ is the previous hidden state,
- *z*_*t*_ is the update gate that controls how much of the previous hidden state is retained,
- *r*_*t*_ is the reset gate that determines how much of the previous hidden state contributes to the candidate activation,
- 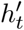 is the candidate activation that combines the current input and the reset-modified previous hidden state,
- *h*_*t*_ is the updated hidden state that blends the previous hidden state and the candidate activation based on the update gate,
- *W*_*\**_ and *U*_*\**_ are trainable weight matrices,
- *b*_*\**_ are bias vectors,
- *σ*(*·*) denotes the sigmoid activation function,
- tanh(*·*) denotes the hyperbolic tangent activation function, and
- *⊙* denotes element-wise multiplication.

The GRU architecture is computationally more efficient than LSTMs due to its reduced complexity, while still maintaining the ability to capture long-term dependencies in sequential data. The update gate allows the model to adaptively control the flow of information from previous time steps, while the reset gate enables the model to forget irrelevant past information when generating the candidate activation. This makes GRUs a popular choice for various sequence modeling tasks, including time series forecasting in epidemiology.

Although LSTMs and GRUs are effective at modeling long-term dependencies, they still rely on sequential processing of data, which can limit their efficiency when dealing with very long sequences. This has led to the development of transformer architectures that leverage self-attention mechanisms to capture dependencies without relying on recurrent connections. Another limitation of RNNs, including LSTMs and GRUs, is that they process sequences sequentially, which can lead to longer training times compared to feedforward models that can be parallelized. This is particularly problematic for long sequences, where the computational cost can become prohibitive. Transformers address this issue by allowing for parallel processing of sequence data, enabling more efficient training and better handling of long-range dependencies.

#### Transformer

The transformer architecture, introduced in [39], is a neural network architecture designed to handle sequential data efficiently that relies on “self-attention” mechanisms. Transformers have revolutionized natural language processing and have been adapted for time series forecasting tasks due to their ability to model long-range dependencies and parallelize computations effectively. The core of a Transformer is self-attention, which lets the model decide how important each time step is when making a prediction. It does this by considering all the data at once and aim to capture the most important parts of the information, assigning weights to all inputs and combining them into a contextual representation. An encoder processes the input sequence, and a decoder uses this information to produce the forecast, while multi-head attention helps capture different types of relationships in the data at the same time, making it particularly effective for complex time series forecasting tasks where multiple factors may influence the target variable.

## B Statistical model

### B.1 ARIMA models

An ARIMA(*p, d, q*) model consists of the following components:

1. AR(*p*): the autoregressive component of order *p*, in which the current observation depends linearly on its previous values.
2. I(*d*): the integrated component of order *d*, representing the number of differencing operations applied to achieve stationarity.
3. MA(*q*): the moving average component of order *q*, in which the current observation depends linearly on previous forecast errors.

Let *{y*_*t*_*}* denote a time series. The differenced series of order *d* is recursively defined as

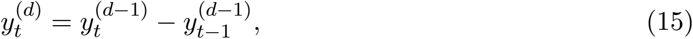

with first-order differencing given by

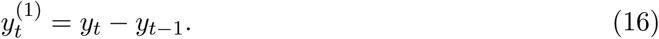

After differencing, the ARIMA(*p, d, q*) model can be expressed as

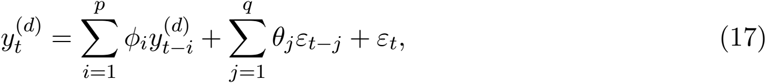

where *ϕ*_*i*_ and *θ*_*j*_ denote the autoregressive and moving average coefficients, respectively, and *ε*_*t*_ represents a white-noise error term.

### B.2 Prophet model

Prophet is a time series forecasting model developed for handling time series with multiple seasonal patterns, trend changes, and missing observations. The model adopts a decomposable framework in which the observed time series is represented as the sum of trend, seasonal, and holiday components. Prophet is particularly suitable for forecasting problems involving strong seasonality and nonlinear trends while maintaining interpretability and computational efficiency.

A brief description of the Prophet model is as follows

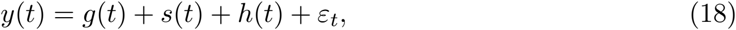

where *g*(*t*) denotes the trend component capturing non-periodic changes in the time series (typically using piecewise linear or logistic growth model), *s*(*t*) represents the seasonal component modeling periodic fluctuations using Fourier series, and *h*(*t*) accounts for the effects of holidays or irregular events. The residual term *ε*_*t*_ captures unexplained variability and random noise.

Prophet has gained popularity on real-world forecasting tasksas it can effectively handle missing data, outliers, and multiple seasonalities without extensive manual preprocessing.

## C Global Forecasting Methodology

The global forecasting methodology follows the cross-learning (or data pooling) paradigm, in which a single forecasting model is trained on multiple related time series. In this study, the global models leverage information from surveillance data collected across 8 Canadian provinces and three respiratory viruses (influenza A, Covid-19, and RSV). Unlike local forecasting, where each model is trained independently for a single province and target virus, global models exploit shared temporal patterns and cross-series dependencies among different provinces and viruses to improve predictive performance.

### Data preparation

Each time series is independently normalized using either Min-Max scaling or standardization. The normalized series is then transformed into a supervised learning dataset via lag embedding, in which lagged observations serve as predictor variables and the subsequent observation is the target variable. The lag-embedded matrices from all selected time series are vertically concatenated to form a single large training matrix. Consequently, model training reduces to a standard supervised regression problem in which the predictor matrix consists of the lagged observations and the response vector contains the corresponding target values (see [73]).

Data splitting, model architecture, hyperparameter search space, and hyperparameter tuning follow the same procedures as in the local paradigm.

### Models

The following global forecasting models are evaluated: LSTM, GRU, and transformer.

## Data Availability

https://www.canada.ca/en/public-health/services/surveillance/respiratory-virus-detections-canada.html

## Ethics statement

Because this study relies exclusively on the secondary use of anonymous information, and the analysis does not generate identifiable information, it did not require a research ethics board review in accordance with Article 2.4 of the Tri-Council Policy Statement: Ethical Conduct for Research Involving Humans (TCPS 2).

## Data accessibility

All clinical laboratory test data used in this study is accessible on the respiratory virus detection portal from Public Health Agency of Canada https://www.canada.ca/en/public-health/services/surveillance/respiratory-virus-detections-canada.html.

## Author contributions

Conceptualization, DC, MC; Data curation, DC, WY; Formal analysis, DC, WY, BBL; Funding acquisition, MC; Methodology, DC, WY, BBL, MC; Software, BBL, WY; Supervision, DC; Validation, DC, WY; Visualization, WY, BBL; Writing – original draft, WY, DC, BBL; Writing – review and editing, WY, DC.

## Acknowledgements

None.

## Disclaimer

None.

## References

[1] World Health Organization et al. Global report on infection prevention and control. World Health Organization, 2022.

[2] Simon Pollett et al. “Identification and evaluation of epidemic prediction and forecasting reporting guidelines: A systematic review and a call for action”. In: Epidemics 33 (2020), p. 100400.

[3] William Ogilvy Kermack and Anderson G McKendrick. “A contribution to the mathematical theory of epidemics”. In: Proceedings of the royal society of london. Series A, Containing papers of a mathematical and physical character 115.772 (1927), pp. 700–721.

[4] Alexander Rodriguez et al. “Machine learning for data-centric epidemic forecasting”. In: Nature Machine Intelligence 6.10 (2024), pp. 1122–1131.

[5] Y Ye et al. Integrating artificial intelligence with mechanistic epidemiological modeling: a scoping review of opportunities and challenges. Nat Commun. 2025; 16 (1): 581.

[6] William Ogilvy Kermack and A. G. McKendrick. “A contribution to the mathematical theory of epidemics”. In: Proceedings of the Royal Society of London. Series A, Containing Papers of a Mathematical and Physical Character 115.772 (1927), pp. 700–721. doi: 10.1098/rspa.1927.0118.

[7] David Champredon, Jonathan Dushoff, and David J. D. Earn. “Equivalence of the Erlang-Distributed SEIR Epidemic Model and the Renewal Equation”. In: SIAM Journal on Applied Mathematics 78.6 (2018), pp. 3258–3278. doi: 10.1137/18M1186411. eprint: https://doi.org/10.1137/18M1186411. url: https://doi.org/10.1137/18M1186411.

[8] Ghazaleh Babanejaddehaki, Aijun An, and Manos Papagelis. “Disease outbreak detection and forecasting: A review of methods and data sources”. In: ACM Transactions on Computing for Healthcare 6.2 (2025), pp. 1–40.

[9] Yiming Fei. “Analysis the use of machine learning algorithm-based methods in predicting COVID-19 infection”. In: Proceedings of the 2nd International Symposium on Artificial Intelligence for Medicine Sciences. 2021, pp. 88–94.

[10] Pavan Kumar et al. “Forecasting the dynamics of COVID-19 pandemic in top 15 countries in April 2020: ARIMA model with machine learning approach”. In: MedRxiv (2020), pp. 2020–03.

[11] Saleh I Alzahrani, Ibrahim A Aljamaan, and Ebrahim A Al-Fakih. “Forecasting the spread of the COVID-19 pandemic in Saudi Arabia using ARIMA prediction model under current public health interventions”. In: Journal of infection and public health 13.7 (2020), pp. 914–919.

[12] KE ArunKumar et al. “Forecasting the dynamics of cumulative COVID-19 cases (con-firmed, recovered and deaths) for top-16 countries using statistical machine learning models: Auto-Regressive Integrated Moving Average (ARIMA) and Seasonal Auto-Regressive Integrated Moving Average (SARIMA)”. In: Applied soft computing 103 (2021), p. 107161.

[13] Santanu Roy, Gouri Sankar Bhunia, and Pravat Kumar Shit. “Spatial prediction of COVID-19 epidemic using ARIMA techniques in India”. In: Modeling earth systems and environment 7.2 (2021), pp. 1385–1391.

[14] Raydonal Ospina et al. “An overview of forecast analysis with ARIMA models during the COVID-19 pandemic: Methodology and case study in Brazil”. In: Mathematics 11.14 (2023), p. 3069.

[15] George Box and GM Jenkins. “Analysis: Forecasting and control”. In: San francisco 10 (1976).

[16] Sean J Taylor and Benjamin Letham. “Forecasting at scale”. In: The American Statistician 72.1 (2018), pp. 37–45.

[17] Domenico Benvenuto et al. “Application of the ARIMA model on the COVID-2019 epidemic dataset”. In: Data in brief 29 (2020), p. 105340.

[18] Nalini Chintalapudi, Gopi Battineni, and Francesco Amenta. “COVID-19 virus outbreak forecasting of registered and recovered cases after sixty day lockdown in Italy: A data driven model approach”. In: Journal of Microbiology, Immunology and Infection 53.3 (2020), pp. 396–403.

[19] Zeynep Ceylan. “Estimation of COVID-19 prevalence in Italy, Spain, and France”. In: Science of The Total Environment 729 (2020), p. 138817.

[20] Qingquan Chen et al. “Prediction of influenza outbreaks in Fuzhou, China: comparative analysis of forecasting models”. In: BMC Public Health 24.1 (2024), p. 1399.

[21] Suprabhath Kalahasti et al. “Foundation models for time series forecasting and policy evaluation in infectious disease epidemics”. In: Epidemics (2026), p. 100916.

[22] Gerardo Chowell. “Fitting dynamic models to epidemic outbreaks with quantified uncertainty: A primer for parameter uncertainty, identifiability, and forecasts”. In: Infectious Disease Modelling 2.3 (2017), pp. 379–398.

[23] Nicholas G Reich et al. “A collaborative multiyear, multimodel assessment of seasonal influenza forecasting in the United States”. In: Proceedings of the National Academy of Sciences 116.8 (2019), pp. 3146–3154.

[24] Arthur L Samuel. “Some studies in machine learning using the game of checkers”. In: IBM Journal of research and development 3.3 (1959), pp. 210–229.

[25] Samuel Dixon et al. “A comparison of infectious disease forecasting methods across locations, diseases, and time”. In: Pathogens 11.2 (2022), p. 185.

[26] Omer Sagi and Lior Rokach. “Ensemble learning: A survey”. In: Wiley interdisciplinary reviews: data mining and knowledge discovery 8.4 (2018), e1249.

[27] Mudasir A Ganaie et al. “Ensemble deep learning: A review”. In: Engineering applications of artificial intelligence 115 (2022), p. 105151.

[28] Martin S Wohlfender et al. “Machine learning-based short-term forecasting of COVID-19 hospital admissions using routine hospital patient data”. In: Epidemics (2025), p. 100877.

[29] Ravid Shwartz-Ziv and Amitai Armon. “Tabular data: Deep learning is not all you need”. In: Information fusion 81 (2022), pp. 84–90.

[30] Trevor Hastie. “The elements of statistical learning data mining, inference, and prediction”. In: (2005).

[31] Martin S. Wohlfender et al. “Machine learning-based short-term forecasting of COVID-19 hospital admissions using routine hospital patient data”. In: Epidemics 54 (2026), p. 100877. issn: 1755-4365. doi: 10.1016/j.epidem.2025.100877. url: https://www.sciencedirect.com/science/article/pii/S1755436525000659.

[32] Shereen Elsayed et al. “Do we really need deep learning models for time series forecasting?” In: arXiv preprint arXiv:2101.02118 (2021).

[33] Angelo Casolaro et al. “Deep learning for time series forecasting: Advances and open problems”. In: Information 14.11 (2023), p. 598.

[34] Rohitash Chandra, Shaurya Goyal, and Rishabh Gupta. “Evaluation of deep learning models for multi-step ahead time series prediction”. In: Ieee Access 9 (2021), pp. 83105–83123.

[35] Ping Jiang. “Nowcasting influenza using Google flu trend and deep learning model”. In: 2020 2nd International Conference on Economic Management and Cultural Industry (ICEMCI 2020). Atlantis Press. 2020, pp. 407–416.

[36] Abdelhafid Zeroual et al. “Deep learning methods for forecasting COVID-19 time-Series data: A Comparative study”. In: Chaos, solitons & fractals 140 (2020), p. 110121.

[37] Sepp Hochreiter and Jürgen Schmidhuber. “Long short-term memory”. In: Neural computation 9.8 (1997), pp. 1735–1780.

[38] Kyunghyun Cho et al. “On the properties of neural machine translation: Encoder–decoder approaches”. In: Proceedings of SSST-8, eighth workshop on syntax, semantics and structure in statistical translation. 2014, pp. 103–111.

[39] Ashish Vaswani et al. “Attention is all you need”. In: Advances in neural information processing systems 30 (2017).

[40] Oleksandr Kamyshnyi et al. “Forecasting influenza epidemics and pandemics in the age of AI and machine learning”. In: Reviews in Medical Virology 36.1 (2026), e70107.

[41] Siva Rama Krishna Kottapalli et al. “Foundation Models for Time Series: A Survey”. In: arXiv e-prints (2025), arXiv–2504.

[42] Yuxuan Liang et al. “Foundation models for time series analysis: A tutorial and survey”. In: Proceedings of the 30th ACM SIGKDD conference on knowledge discovery and data mining. 2024, pp. 6555–6565.

[43] Steffen Albrecht et al. “Forecasting severe respiratory disease hospitalizations using machine learning algorithms”. In: BMC Medical Informatics and Decision Making 24.1 (2024), p. 293.

[44] Lucas Moreira Ribeiro, Eduardo Jose da Silva Luz, and Jadson Castro Gertrudes. “How Effective Are Time Series Foundation Models for Epidemiological Data Analysis?” In: 2025 IEEE EMBS International Conference on Biomedical and Health Informatics (BHI). IEEE. 2025, pp. 1–7.

[45] Yanqin Wang et al. “A comparative study of time series foundation models for hand, foot, and mouth disease forecasting: TimesFM, Moirai, and traditional approaches”. In: Frontiers in Public Health 13 (2025), p. 1634138.

[46] Abdul Fatir Ansari et al. “Chronos: Learning the language of time series”. In: arXiv preprint arXiv:2403.07815 (2024).

[47] Abhimanyu Das et al. “A decoder-only foundation model for time-series forecasting”. In: arXiv preprint arXiv:2310.10688 (2023).

[48] Sarah Berkhahn and Matthias Ehrhardt. “A physics-informed neural network to model COVID-19 infection and hospitalization scenarios”. In: Advances in continuous and discrete models 2022.1 (2022), p. 61.

[49] Caterina Millevoi, Damiano Pasetto, and Massimiliano Ferronato. “A physics-informed neural network approach for compartmental epidemiological models”. In: PLOS Computational Biology 20.9 (2024), e1012387.

[50] Yulia Abramova and Vasiliy Leonenko. “The past helps the future: coupling differential equations with machine learning methods to model epidemic outbreaks”. In: International Conference on Computational Science. Springer. 2024, pp. 247–254.

[51] Jiwen Xia et al. “Physics-Informed Neural Networks for Infectious Disease Modeling with Limited Data”. In: 2024 IEEE/WIC International Conference on Web Intelligence and Intelligent Agent Technology (WI-IAT). IEEE. 2024, pp. 508–513.

[52] Ying Qian et al. “Physics-informed deep learning for infectious disease forecasting”. In: Arxiv (2025), arXiv–2501.

[53] Martina Rama et al. “Forecasting seasonal influenza epidemics with physics-informed neural networks”. In: Epidemics (2026), p. 100919.

[54] Achraf Zinihi, Matthias Ehrhardt, and Moulay Rchid Sidi Ammi. “A Koopman-PINN Framework for Epidemic Models: Parameter Inference and Forecasting”. In: arXiv preprint arXiv:2606.15201 (2026).

[55] Moritz UG Kraemer et al. “Artificial intelligence for modelling infectious disease epidemics”. In: Nature 638.8051 (2025), pp. 623–635.

[56] Hassan Ali. “AI for pandemic preparedness and infectious disease surveillance: predicting outbreaks, modeling transmission, and optimizing public health interventions”. In: Int J Res Publ Rev 5.8 (2024), pp. 4605–19.

[57] Yang Ye et al. “Integrating artificial intelligence with mechanistic epidemiological modeling: a scoping review of opportunities and challenges”. In: Nature Communications 16.1 (2025), p. 581.

[58] Hooman H Rashidi et al. “Introduction to artificial intelligence and machine learning in pathology and medicine: generative and nongenerative artificial intelligence basics”. In: Modern Pathology 38.4 (2025), p. 100688.

[59] Neha Sharma, Reecha Sharma, and Neeru Jindal. “Machine learning and deep learning applications-a vision”. In: Global Transitions Proceedings 2.1 (2021), pp. 24–28.

[60] Alexander Rodríguez, Harshavardhan Kamarthi, and B Aditya Prakash. “Epidemic Fore-casting with a Data-Centric Lens”. In: Proceedings of the 28th ACM SIGKDD Conference on Knowledge Discovery and Data Mining. 2022, pp. 4822–4823.

[61] Xue Bai and Yang Yu. “Research and Application of Time Series Forecasting Methods in Disease Surveillance”. In: IJLAI Transactions on Science and Engineering 4.1 (2026), pp. 1–10.

[62] Omar Enzo Santangelo et al. “Machine learning and prediction of infectious diseases: a systematic review”. In: Machine Learning and Knowledge Extraction 5.1 (2023), pp. 175–198.

[63] Matheus Henrique Dal Molin Ribeiro et al. “Short-term forecasting COVID-19 cumulative confirmed cases: Perspectives for Brazil”. In: Chaos, Solitons & Fractals 135 (2020), p. 109853.

[64] Vinay Kumar Reddy Chimmula and Lei Zhang. “Time series forecasting of COVID-19 transmission in Canada using LSTM networks”. In: Chaos, solitons & fractals 135 (2020), p. 109864.

[65] Alfred B Amendolara et al. “LSTM-based recurrent neural network provides effective short term flu forecasting”. In: BMC Public Health 23.1 (2023), p. 1788.

[66] Daiki Koge and Keita Wagatsuma. “Long short-term memory-based forecasting of influenza epidemics using surveillance and meteorological data in Tokyo, Japan”. In: Frontiers in Public Health 13 (2025), p. 1618508.

[67] Neo Wu et al. “Deep transformer models for time series forecasting: The influenza prevalence case”. In: arXiv preprint arXiv:2001.08317 (2020).

[68] Edmund Fosu Agyemang, Hansapani Rodrigo, and Vincent Agbenyeavu. “Comparative analysis of traditional and deep learning time series architectures for influenza A infectious disease forecasting”. In: Computers in Biology and Medicine 211 (2026), p. 111736.

[69] Edmund F Agyemang, Hansapani Rodrigo, and Vincent Agbenyeavu. “A Comparative Analysis of Traditional and Deep Learning Time Series Architectures for Influenza A Infectious Disease Forecasting”. In: arXiv preprint arXiv:2507.19515 (2025).

[70] Farah Shahid, Aneela Zameer, and Muhammad Muneeb. “Predictions for COVID-19 with deep learning models of LSTM, GRU and Bi-LSTM”. In: Chaos, Solitons & Fractals 140 (2020), p. 110212.

[71] Noah Hollmann et al. “Tabpfn: A transformer that solves small tabular classification problems in a second”. In: arXiv preprint arXiv:2207.01848 (2022).

[72] Azul Garza, Cristian Challu, and Max Mergenthaler-Canseco. “TimeGPT-1”. In: arXiv preprint arXiv:2310.03589 (2023).

[73] Pablo Montero-Manso and Rob J Hyndman. “Principles and algorithms for forecasting groups of time series: Locality and globality”. In: International Journal of Forecasting 37.4 (2021), pp. 1632–1653.

[74] Takuya Akiba et al. “Optuna: A next-generation hyperparameter optimization frame-work”. In: Proceedings of the 25th ACM SIGKDD international conference on knowledge discovery & data mining. 2019, pp. 2623–2631.

[75] Diederik P Kingma and Jimmy Ba. “Adam: A method for stochastic optimization”. In: arXiv preprint arXiv:1412.6980 (2014).

[76] David Champredon and Jonathan Dushoff. “Intrinsic and realized generation intervals in infectious-disease transmission”. In: Proceedings of the Royal Society B: Biological Sciences 282.1821 (Dec. 2015), p. 20152026. issn: 0962-8452. doi: 10.1098/rspb.2015.2026. eprint: https://royalsocietypublishing.org/rspb/article-pdf/doi/10.1098/rspb.2015.2026/607706/rspb.2015.2026.pdf. url: https://doi.org/10.1098/rspb.2015.2026.

[77] Angel N Desai et al. “Real-time epidemic forecasting: challenges and opportunities”. In: Health security 17.4 (2019), pp. 268–275.

[78] Dianbo Liu et al. “Real-time forecasting of the COVID-19 outbreak in Chinese provinces: Machine learning approach using novel digital data and estimates from mechanistic models”. In: Journal of medical Internet research 22.8 (2020), e20285.

[79] Yann LeCun, Yoshua Bengio, and Geoffrey Hinton. “Deep learning”. In: nature 521.7553 (2015), pp. 436–444.

